# Predicting highly pathogenic avian influenza H5N1 outbreak risk using extreme weather and bird migration data in machine learning models

**DOI:** 10.64898/2026.03.30.26349797

**Authors:** William W. Zou, Elizabeth J. Carlton, Elise N. Grover

## Abstract

**Background:** Climate change is intensifying extreme weather events (EWEs) with potentially profound consequences for zoonotic disease dynamics, yet the mechanisms linking EWEs to highly pathogenic avian influenza (HPAI) H5N1 outbreaks remain poorly characterized. The ongoing H5N1 panzootic — responsible for infection in over 500 avian and mammalian species, as well as nearly 1000 human cases and 477 deaths worldwide — provides a critical opportunity to evaluate how climate conditions shape spillover risk at landscape scales.

**Methods:** We compiled a county-month dataset of confirmed H5N1 detections across the contiguous United States from 2022–2024 and integrated it with satellite-derived climate metrics, storm event data, and wild bird activity data. We trained and validated a gradient boosting machine classifier to predict outbreak risk and characterize predictor relationships.

**Results:** Our model achieved strong discriminative performance (AUC-ROC = 0.856; AUC-PR = 0.237, representing a 7-fold improvement over chance) and high recall (0.726), supporting its utility as an early warning tool. Human population and temperature-related variables were the most influential predictors: cold temperature shocks and prolonged low temperatures were consistently associated with elevated outbreak risk, likely through enhanced environmental viral persistence, wild bird habitat compression, and allostatic stress-driven immunosuppression in reservoir hosts. Among storm variables, high wind coverage elevated risk, potentially via aerosol dispersal of contaminated particulates, while tornado activity showed an inverse relationship, consistent with documented avoidant behavior in migratory birds. Wild bird reservoir density showed a strong positive monotonic relationship with outbreak risk.

**Conclusions:** Our analyses demonstrate that routinely available environmental and infection data can be used to predict HPAI outbreak risk at fine spatiotemporal scales. These findings demonstrate the divergent roles of short-versus long-term environmental exposures in HPAI spillover dynamics, as well as the potential for machine learning-based surveillance tools to inform targeted biosecurity interventions and early warning systems.

## 1. Introduction

Climate change is reshaping the global landscape of infectious diseases (1–3) by driving massive shifts in temperature, precipitation, and the frequency and intensity of Extreme Weather Events (EWEs) such as tornadoes, floods and wildfires (4). These climate forces can transform reservoir host distribution and patterns of contact at the wildlife–agriculture–human interface, influencing where, when, and how often disease spillover occurs (5–7). Within this context, Highly Pathogenic Avian Influenza (HPAI), particularly H5N1, has emerged as a pathogen of global concern (8). H5N1 is an HPAI virus primarily maintained in migratory waterfowl, which serves as its natural reservoir and drives its global spread (9, 10). Since its initial emergence in Scotland in 1959 (11), the virus has caused mortality in wild birds and large-scale outbreaks in domestic poultry through spillovers from infected migratory populations (12). Recurrent and increasing spillover events have also been documented across numerous mammal species, including sporadic infections among human populations that are primarily linked to occupational exposures (13–15). As of March 2026, there have been 993 reported human cases of H5N1 across 25 countries, with a 48% case-fatality rate (477 deaths) worldwide (16).

Despite its initial emergence in 1959, HPAI H5N1 was only first detected in wild bird populations in North America in November 2021 (18). Since then, there have been over 10,000 cases from more than 160 North American wild bird species confirmed to date, and evidence of spillover to over 400 wild mammals from over 20 distinct species (19). The current North American HPAI wave has led to 71 human infections (20), one death (21), and the loss of over 100 million poultry (22). While human infections are currently episodic and most typically characterized as mild (23), the risk of HPAI viruses developing sustained human-to-human transmission is heightened by ongoing genetic reassortment and cross-species transmission (24, 25). A new circulating genotype of H5N1 belonging to clade 2.3.4.4b, first detected in Europe in 2021, is primarily responsible for crossing species barriers in North America, infecting a growing range of mammals, including dairy cattle, cats and sea lions (17, 26). This expanding host range is especially concerning given influenza A’s segmented genome (12), which allows different viral genotypes, for example, a mammal-adapted lineage such as B3.13 (27) and an avian-adapted lineage such as D1.1 (28), to exchange genetic material through reassortment, raising the risk of emergent variants that combine efficient human infectivity with high virulence (17). Given the expanding geographic distribution (29) and increasing frequency of spillovers (30), we see an urgent need to understand the environmental and climatic patterns that are shaping local H5N1 spillover events and transmission potential.

Climate, generally, and EWEs in particular, may play an important but yet poorly documented role in H5N1 outbreaks in poultry. Environmental conditions can shape both host movement and pathogen persistence, the two primary drivers of spillover risk. EWEs such as droughts, heat waves, wildfires, and hurricanes can disrupt the behavioral ecology of migratory waterfowl while simultaneously altering the environmental conditions that govern viral survival and dispersal (31). These dual forces inform our definition and selection of predictors to capture shifts in climate and EWEs that we hypothesize can alter host movement and pathogen persistence, and by extension, the overall transmission risk (see Supplementary Table S1).

At the host level, EWEs can alter migration timing, displacement routes, and stopover site selection in migratory waterfowl (32). For example, the desiccation of inland wetlands during prolonged drought has been linked to higher concentrations of birds and mammals around remaining water bodies (33), increasing contact rates with domestic poultry and the likelihood of spillover events. Similarly, wildfires have been linked to large-scale displacement and disrupted foraging behavior (34), producing comparable increases in host-level transmission risk.

At the virus level, temperature and precipitation shocks can affect viral persistence and dispersal in water and sediments (35–37), influencing how long infectious particles remain viable in the environment. These mechanisms suggest that a range of environmental pressures can alter infectious disease transmission dynamics at both the host and pathogen levels, making it essential to investigate the relationship between EWE events and HPAI outbreak risk.

Despite this possibility, few studies have directly examined how EWEs shape HPAI outbreak risk across space and time. The recent incursion and establishment of H5N1 in North America provides an opportunity to test these relationships.

Here, we address this critical gap by evaluating the relationships between EWEs and H5N1 outbreaks using machine learning models. We hypothesize that certain EWEs, such as wildfires and drought, may reduce outbreak risk by deflecting migration routes away from affected regions, whereas others such as high precipitation, flooding and colder-than-normal temperatures may increase risk by creating favorable conditions for stopovers, redirecting birds into high-density poultry zones or promoting viral stability. By combining disease surveillance, ecological and climate data, with machine learning methods, we aimed to identify important environmental predictors of H5N1 outbreaks and provide interpretable relationships.

## 2. Methods

### 2.1 Outbreak Data

To investigate the role and importance of EWEs in H5N1 outbreaks, we compiled a three-year dataset (2022–2024) of confirmed H5N1 detections in commercial poultry, backyard flocks and wild bird populations at a county-month resolution across the contiguous United States and integrated it with satellite-derived climate metrics and crowd-sourced migration data for use in Gradient Boosting Machine (GBM) prediction models.

Outbreak data were obtained from the US Department of Agriculture’s (USDA) Animal and Plant Health Inspection Service (APHIS) (38). APHIS reports all individual detections of H5N1 within commercial poultry, backyard flock and wild bird populations in the US. We defined an outbreak as the detection of one or more cases within a flock between January 2022 to December 2024. We aggregated outbreaks to a county–month scale and converted them to a binary outcome indicating the presence or absence of outbreaks each month within each US county.

### 2.2 Extreme Weather Events Data

We included two types of EWEs as model predictors: 1) weather conditions and 2) storm events.

Weather conditions included county-level monthly average surface temperature, total precipitation, and drought and hydrology (as measured by the Palmer Drought Severity Index (PDSI) and the Palmer Hydrological Drought Index (PHDI), respectively) (39, 40), each of which were retrieved from the National Oceanic Atmospheric Association’s (NOAA) database with an automated scraping process. We constructed two classes of predictors for each included weather condition (Table S1). The first class captured recent, baseline weather trends using a 90-day rolling mean, calculated as the mean of the three months prior to month *t*:

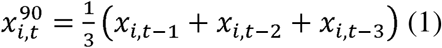

Where *x_i,t-k_* denotes the value of predictor *x* in county *i*, k months prior to prediction month *t*. This measure is intended to capture sustained environmental conditions such as prolonged drought or persistent temperatures that may gradually alter host movement patterns and pathogen persistence over the months preceding an outbreak. The second class captured weather shocks, defined as the deviation of the most recent month’s conditions from the preceding 90-day trend:

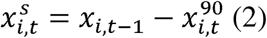

Where 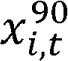 is the rolling mean defined in equation (1). This shock measure is designed to capture abrupt departures from recent baseline conditions that may acutely disrupt host behavior or alter pathogen persistence. Although *x_i,t-l_* appears in both the shock measure and the rolling mean, we retain it in the baseline calculation because a three-month window best captures the recent environmental trend against which weather shocks should be assessed; excluding *x_i,t-l_* from the mean would produce a baseline anchored further in the past, potentially conflating true shocks with lagged seasonal transitions rather than isolated departures from the most recent local conditions. Together, the rolling mean and shock predictors are intended to separately identify the effects of prolonged environmental trends and acute shocks on outbreak risk. For both predictor classes, we excluded values from the prediction month *t* itself to prevent temporal leakage.

Storm predictors were designed to capture recent county-level storm intensity, sustained storm conditions, and the spatial extent of disruption in the surrounding state. Mechanistically, storms can influence outbreaks by displacing wild birds and altering contact patterns, and by disrupting farm operations, biosecurity, logistics, and surveillance/reporting, thereby changing poultry exposure and the probability that infections are detected. Storm events included dust storms, high winds, floods, hurricanes, tornadoes, and wildfires (Table S1). Data on county storm events was retrieved from the NOAA “Storm Events Database” (41). Although the data retrieved was in tabular form, there was limited quantitative data, and most information was provided via narrative reports. As such, we used rule-based string extraction to quantify the severity of the event (a detailed explanation of our string extraction methods may be found in Supplementary Methods S1). We constructed three classes for each storm predictor (dust storms, floods, hurricanes, tornadoes and wildfires). For our first two predictor classes, an inverse hyperbolic sine (asinh) transformation, defined as 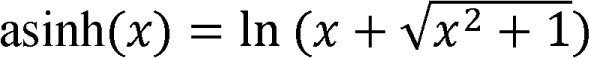, was used because the storm event predictors behave similarly to a log transformation at large values — compressing the right tail of skewed distributions, while remaining defined at zero — making it well-suited for zero-inflated count variables such as storm event frequency at the county-month scale. As such, our first class was a lagged (month *t-1*) asinh-transformed storm predictor, while our second class was an asinh-transformed 90-day rolling mean of storm severity (months *t–1, t-2, t-3*). Our third storm predictor class was designed to capture the effect that statewide neighboring storm events have on a given county. For brevity’s sake, we refer to this class of predictors as the “area affected” by each given storm event. Area affected storm variables were calculated as the lagged (month *t-1*) percent of all counties across the state that experienced the specific storm event, excluding the focal county to eliminate issues with multicollinearity:

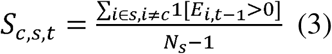

where *S_c,S,t_* is the statewide neighboring storm event measure for focal county c in state s at prediction month *t*, 11[*E*_*j,t*-1_ > 0] is an indicator function equal to 1 if county *j* experience the storm event in month *t-1*, *N_S_* is the total number of counties in state *s*, and the denominator *N_S_* -1 reflects the exclusion of the focal county *c* from the calculation.

### 2.3 Bird Migration Data

We retrieved bird migration data from the North American Bird Banding Program Dataset from the US Geological Survey’s Bird Banding Lab (BBL) (42). This dataset was split into groups containing multiple species of birds. We identified and retrieved groups containing the probable host species for H5N1 from the BBL dataset, including domestic and wild birds based on the Memorial Sloan Kettering Cancer Center’s list of “Affected Bird Species” (Supplementary Table S2) (43). Our final list included 467 bird species. Because bird banding records are generated by a distributed network of volunteer and licensed banders, raw band counts are inherently subject to spatial variation in sampling effort; counties with larger human populations are likely to have more bird banders and thus produce more records independent of true bird activity. To correct for this sampling bias, we normalized unique band counts by county human population using 5-year population estimates from the 2023 American Community Survey (44), an approach consistent with the literature on opportunistic biodiversity data, which commonly uses human population density or accessibility as covariates to account for uneven observer effort across space (45).

Finally, we included county-level human population (log-transformed) to capture variation in factors that scale with population size, including agricultural production intensity, human-wildlife contact rates, and veterinary and public health surveillance infrastructure — all of which can influence both true outbreak risk and the probability that infections are detected and reported.

### 2.4 Interpretable Gradient Boosting Machine Construction

To evaluate the relationship between a range of EWE predictors and H5N1 outbreaks in the US between 2022–2024, we leveraged GBMs, a robust machine learning prediction method often used to study infectious disease outbreaks due to their high capacity for capturing nonlinear effects and high-order interactions while maintaining interpretability and strong predictive performance (46).

To reduce multicollinearity among the 62 candidate predictors, we performed correlation-based clustering using a graph representation of pairwise correlations. We first imported a table of pairwise correlations (Supplementary Figure S1) and applied an absolute-correlation threshold of 0.70. Variables were represented as nodes in an undirected graph, and an edge was drawn between two nodes if their pairwise absolute correlation exceeded 0.70. Correlated variable groups were then defined as connected components of this graph (i.e., sets of variables linked directly or indirectly through above-threshold correlations). For each connected component, we selected a single representative variable to retain. The representative was defined as the variable with the highest degree within its component (i.e., the greatest number of above-threshold correlations to other variables in the same component). In the case of ties, we selected the variable with the highest mean absolute correlation to other variables within the component. Variables that did not exceed the correlation threshold with any other variable formed singleton components and were retained automatically. This procedure yielded a final reduced feature set containing 28 predictors, each representing a single variable per correlation cluster.

All iterations of our GBM models assessed the relationship between our final set of predictors and a binary metric indicating the presence of one or more H5N1 outbreaks at the county-month level. Our models also included categorical indicators of calendar-month, county, and state to absorb fixed heterogeneity and seasonality effects.

The XGBoost package in R (47) was used to train all models, splitting our data into training (80%) and test (20%) sets that were stratified by outbreak status. We optimized model hyperparameters using Optuna, an automated hyperparameter optimization framework that efficiently explores the parameter space by iteratively proposing candidate configurations, evaluating them, and updating the search strategy based on prior results. For each Optuna trial, we performed 5-fold stratified cross-validation on the training set and used the mean Area Under the Precision-Recall Curve (AUC-PR) across folds as the objective to maximize. We optimized hyperparameters by maximizing AUC-PR because precision–recall performance is more informative than the Receiver Operating Characteristic AUC (ROC-AUC) when positive outcomes are rare, as AUC-PR directly reflects the performance of identifying positives (48). Tuned hyperparameters included tree count, depth, learning rate, subsampling, column subsampling, and regularization terms. Because outbreaks were rare, we applied class weighting in each cross-validation fold. We computed a base positive-class weight as the ratio of negative to positive samples in the training fold and multiplied this by a tuned scalar. After selecting optimal hyperparameters, we refit the final model on the full training set using the corresponding final positive-class weight and evaluated performance on the held-out test set using AUC-PR, AUC-ROC, accuracy, precision, recall, and F1 score.

We interpreted our model using two complementary post-hoc explainability outputs (49) derived on the held-out test set: We assessed model interpretability using (i) global Shapley Additive exPlanations (SHAP) feature importance and (ii) partial dependence plots (PDPs) for the top-ranked numeric predictors. SHAP values quantify how each predictor shifts an individual prediction above or below the model’s baseline, and global importance was summarized as mean absolute SHAP (mean|SHAP|) across test observations and displayed as dot-and-whisker plots, where larger values indicate greater average influence on predicted outbreak probability. PDPs summarize the model’s marginal relationship between a predictor and predicted outbreak risk by varying that predictor over its observed range while averaging over the joint distribution of all other covariates, thereby visualizing the direction and nonlinearity of the modeled association.

To quantify uncertainty in model interpretation, we used a bootstrap resampling procedure. For each of 25 replicates, we sampled county–month observations from the training data with replacement, separately within outbreak and non-outbreak strata to preserve class balance, refit the model pipeline on the resampled training set, and then recomputed SHAP importances and PDPs on the unchanged test set. We derived 95% confidence intervals from the 2.5^th^ and 97.5^th^ percentiles of the bootstrap distributions.

## 3. Results

### 3.1 Descriptive Statistics

Between January 2022 and December 2024, 3.27% of county-month observations within the contiguous United States experienced at least one H5N1 outbreak during our study period. A total of 13,163 H5N1 outbreaks were detected across 1,785 US counties, encompassing both wild bird detections (n = 11,690; 88.8%) and commercial or backyard poultry flocks (n = 1,473; 11.2%) (Figure 1). Outbreak activity was highest in 2022 (n = 7,021), declining in subsequent years (2023: n = 3,450; 2024: n = 2,692). Seasonally, outbreaks were most frequent in fall (n = 5,857), followed by winter (n = 3,985), spring (n = 2,340), and summer (n = 981), with wild bird detections driving this pattern across all seasons. Seasonally, outbreaks were most frequent in fall and least frequent in summer; however, poultry outbreaks peaked in winter rather than fall, diverging from the pattern seen in wild birds. At the county level, the median number of cumulative outbreaks was 2, with considerable right skew (mean = 7.4, SD = 17.3) reflecting a small number of heavily affected counties. Among affected poultry flocks, an estimated 162,801,168 birds were impacted over the study period, with a mean flock size of 251,236 birds (median: 150; max: 12,108,000), reflecting the substantial heterogeneity in operation size between commercial and backyard poultry producers.

**Figure 1.**
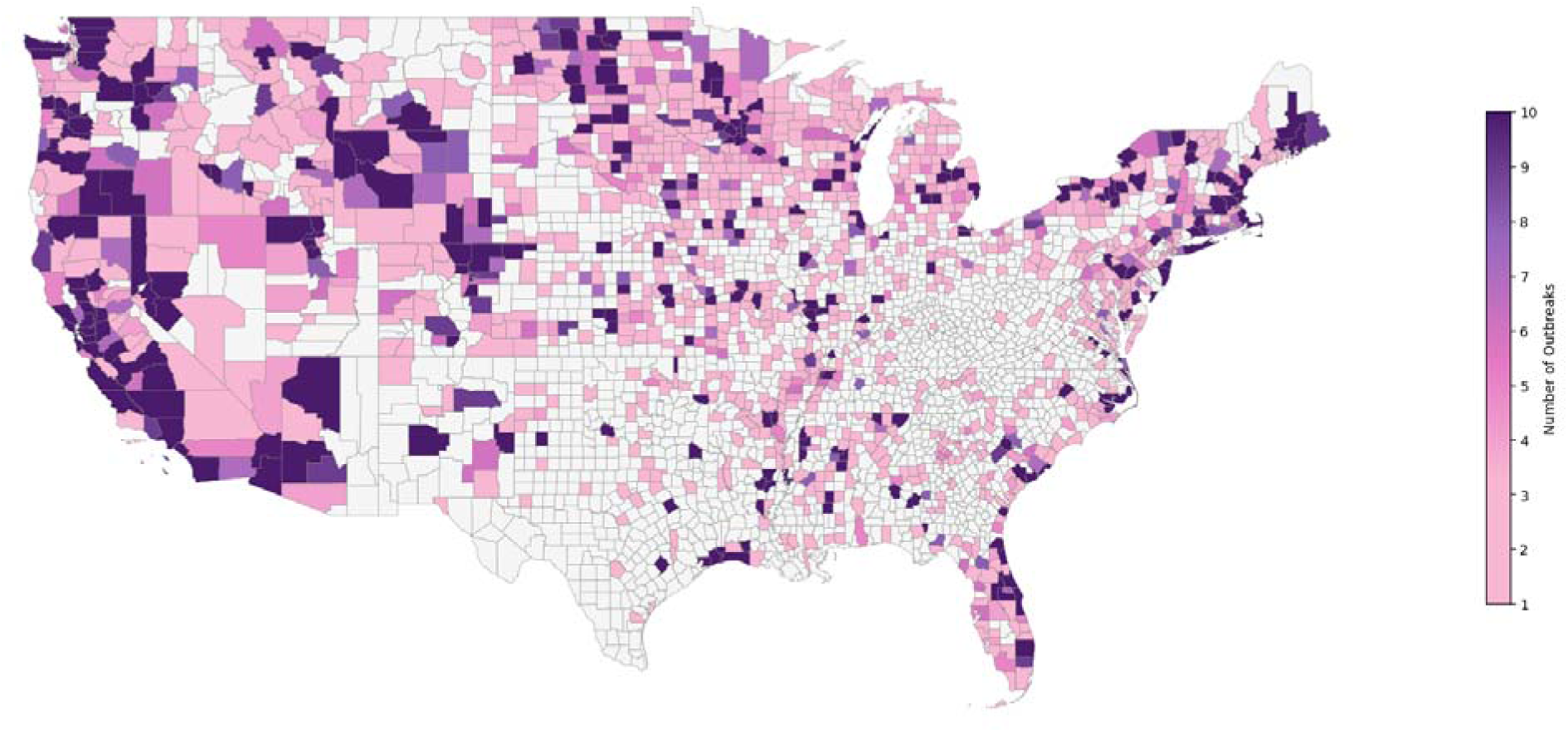
Cumulative HPAI H5N1 outbreaks detected in backyard flocks, commercial poultry and wild birds by county in the contiguous United States, 2022–2024. County-level outbreak counts were derived from USDA APHIS surveillance data. Color intensity reflects the cumulative number of reported outbreaks, ranging from light pink (1 outbreak) to dark purple (≥10 outbreaks). Counties with no reported outbreaks are shown in light gray. County boundaries were obtained from the U.S. Census Bureau TIGER/Line cartographic boundary files (2021). Alaska, Hawaii, and U.S. territories are excluded.

Table 1 presents descriptive statistics for the 28 predictors included in the analysis. Bird reservoir presence was highly right-skewed (Mean = 4.50, SD = 43.59), reflecting the uneven distribution of avian activity or avian activity surveillance across the country. Average 90-day temperatures had an overall mean of 14.61°C (SD = 9.25°C), with the highest average 90-day temperature in the Southern US (Mean = 17.30°C, SD = 7.02°C), and lowest average 90-day temperature in the Western US (Mean = 10.58°C, SD = 8.36°C). Temperature shocks were similar across all US regions. Overall, 90-day precipitation averaged 8.15 mm (SD = 4.37 mm), was highest in the Northeast (Mean = 9.95, SD = 2.74) and lowest in the West (Mean = 4.86, SD = 5.25). Storm events were predominantly near zero in both mean and standard deviation, reflecting the rarity of events such as hurricanes (Mean = 0.00, SD = 0.00 for area affected) and wildfires (Mean = 0.02, SD = 0.32 for asinh-transformed acres), while wind and tornado variables showed slightly greater variation.

**Table 1.** Distribution of 28 county-month predictors by US region, including demographic, weather and storm-related predictors (2022–2024).

| Variable | Northeast<sup>*</sup> |  | Midwest<sup>†</sup> |  | South<sup>‡</sup> |  | West<sup>§</sup> |  |
| --- | --- | --- | --- | --- | --- | --- | --- | --- |
|  | Mean | SD | Mean | SD | Mean | SD | Mean | SD |
| County human population (log-transformed) | 11.64 | 1.27 | 9.93 | 1.42 | 10.33 | 1.35 | 10.30 | 1.85 |
| Bird reservoir presence (log-transformed) | 12.58 | 71.75 | 5.56 | 46.97 | 2.32 | 29.78 | 5.20 | 53.88 |
| Temperature shock (°C) (lag 1 month) | 17.13 | 8.13 | 17.05 | 9.35 | 17.30 | 7.02 | 17.39 | 8.29 |
| Precipitation shock (mm) (lag 1 month) | -0.14 | 5.28 | 0.03 | 5.00 | -0.10 | 6.85 | 0.13 | 5.74 |
| Drought shock (PDSI) (lag 1 month) | -0.09 | 1.29 | -0.06 | 1.29 | 0.01 | 1.12 | 0.08 | 1.40 |
| Hydrology shock (PHDI) (lag 1 month) | -0.13 | 1.28 | -0.05 | 1.27 | -0.02 | 1.10 | 0.09 | 1.29 |
| Avg 90-day temperature (°C) | 11.36 | 7.50 | 11.89 | 8.80 | 18.28 | 6.88 | 10.58 | 8.36 |
| Avg 90-day precipitation (mm) | 9.95 | 2.74 | 6.89 | 3.26 | 9.78 | 4.13 | 4.86 | 5.25 |
| Avg 90-day drought (PDSI) | 0.69 | 1.76 | -0.52 | 2.00 | -0.71 | 1.68 | -0.81 | 2.13 |
| Avg 90-day hydrology (PHDI) | 0.86 | 1.84 | -0.53 | 2.12 | -0.67 | 1.82 | -0.89 | 2.31 |
| Dust magnitude (asinh, lag 1 month) | 0.00 | 0.00 | 0.00 | 0.04 | 0.00 | 0.02 | 0.00 | 0.07 |
| Flood duration (asinh, lag 1 month) | 0.11 | 0.54 | 0.12 | 0.73 | 0.06 | 0.46 | 0.10 | 0.59 |
| High-wind speed (asinh, lag 1 month) | 0.26 | 1.06 | 0.14 | 0.78 | 0.10 | 0.67 | 0.30 | 1.15 |
| Hurricane wind speed (asinh, lag 1 month) | 0.00 | 0.00 | 0.00 | 0.00 | 0.00 | 0.16 | 0.00 | 0.00 |
| Tornado magnitude (asinh, lag 1 month) | 0.09 | 0.74 | 0.18 | 1.06 | 0.20 | 1.17 | 0.05 | 0.48 |
| Wildfire acres (asinh, lag 1 month) | 0.01 | 0.22 | 0.00 | 0.20 | 0.02 | 0.42 | 0.09 | 0.85 |
| Avg 90-day dust magnitude (asinh) | 0.00 | 0.00 | 0.00 | 0.03 | 0.00 | 0.01 | 0.00 | 0.05 |
| Avg 90-day flood duration (asinh) | 0.11 | 0.33 | 0.12 | 0.53 | 0.06 | 0.34 | 0.10 | 0.38 |
| Avg 90-day high-wind speed (asinh) | 0.28 | 0.75 | 0.14 | 0.53 | 0.11 | 0.49 | 0.30 | 0.85 |
| Avg 90-day hurricane wind speed (asinh) | 0.00 | 0.00 | 0.00 | 0.00 | 0.00 | 0.09 | 0.00 | 0.00 |
| Avg 90-day tornado magnitude (asinh) | 0.09 | 0.46 | 0.19 | 0.64 | 0.21 | 0.72 | 0.05 | 0.29 |
| Avg 90-day wildfire acres (asinh) | 0.00 | 0.11 | 0.00 | 0.12 | 0.02 | 0.29 | 0.09 | 0.64 |
| Area affected by dust (lag 1 month) | 0.00 | 0.00 | 0.00 | 0.01 | 0.00 | 0.00 | 0.00 | 0.01 |
| Area affected by flood (lag 1 month) | 0.05 | 0.11 | 0.03 | 0.06 | 0.02 | 0.05 | 0.04 | 0.10 |
| Area affected by high winds (lag 1 month) | 0.06 | 0.13 | 0.03 | 0.07 | 0.02 | 0.05 | 0.06 | 0.09 |
| Area affected by hurricane (lag 1 month) | 0.00 | 0.00 | 0.00 | 0.00 | 0.00 | 0.01 | 0.00 | 0.00 |
| Area affected by tornado (lag 1 month) | 0.02 | 0.04 | 0.04 | 0.06 | 0.03 | 0.06 | 0.01 | 0.03 |
| Area affected by wildfire (lag 1 month) | 0.00 | 0.01 | 0.00 | 0.00 | 0.00 | 0.01 | 0.01 | 0.02 |

<sup>\*</sup> Northeast region includes the following US states: Connecticut, Maine, Massachusetts, New Hampshire, New Jersey, New York, Pennsylvania, Rhode Island, Vermont
<sup>†</sup> Midwest region includes the following US states: Illinois, Indiana, Michigan, Ohio, Wisconsin, Iowa, Kansas, Minnesota, Missouri, Nebraska, North Dakota, South Dakota
<sup>‡</sup> South region includes the following US states: Delaware, District of Columbia, Florida, Georgia, Maryland, North Carolina, South Carolina, Virginia, West Virginia, Alabama, Kentucky, Mississippi, Tennessee, Arkansas, Louisiana, Oklahoma, Texas
<sup>§</sup> West region includes the following US states: Alaska, Arizona, California, Colorado, Hawaii, Idaho, Montana, Nevada, New Mexico, Oregon, Utah, Washington, Wyoming

**Table 2.** The results for 5-fold cross validation with bootstrapped 95% confidence intervals.

3.3 Predictor Importance and Relationship with Outbreak
| Performance Metric | Mean (95% Bootstrap Interval) |
| --- | --- |
| AUC-ROC | 0.856 (0.847-0.864) |
| AUC-PR | 0.237 (0.222-0.250) |
| Accuracy | 0.860 (0.673-0.943) |
| F1 | 0.290 (0.146-0.315) |
| Precision | 0.181 (0.08-0.261) |
| Recall | 0.726 (0.396-0.847) |

### 3.2 Model Performance

Our H5N1 county-month GBM classifier achieved AUC-PR = 0.237 (95% CI: 0.222-0.250) and AUC-ROC = 0.856 (95% CI: 0.847-0.864) on the held-out test set, with an accuracy = 0.860 (95% CI: 0.673-0.943), precision = 0.181 (95% CI: 0.08-0.261), recall = 0.726 (95% CI: 0.396-0.847), and F1 = 0.290 (95% CI: 0.146-0.315) (Table 1). Because outbreaks are rare (3.27% baseline prevalence), precision–recall is the most informative summary: a random classifier would have AUC-PR ≈ 0.033, so our model delivers roughly a 7.18x improvement in average precision over chance while retaining good rank separation by ROC.

### 3.3 Predictor Importance and Relationship with Outbreak

We found that the most important predictors of H5N1 outbreaks based on mean|SHAP| values were county-level human population and temperature shocks (Figure 2). Four out of the top five predictors of H5N1 outbreaks were weather variables, two of which represented precipitation (average 90-day precipitation and precipitation shock) and two represented temperature (average 90-day temperature and temperature shock). The two most important storm predictors were the area affected by tornadoes in the previous month and the area affected by high winds in the previous month. Bird reservoir density also played an important role and was ranked as the sixth most important predictor. Predictors that had a negligible contribution to the predictive performance of the model (mean|SHAP| < 0.01) were area affected by hurricanes, average 90-day acres burned by wildfires, acres burned by wildfires, average 90-day dust storm strength, average 90-day hurricane wind speed, hurricane wind speed, and dust storm strength.

**Figure 2.**
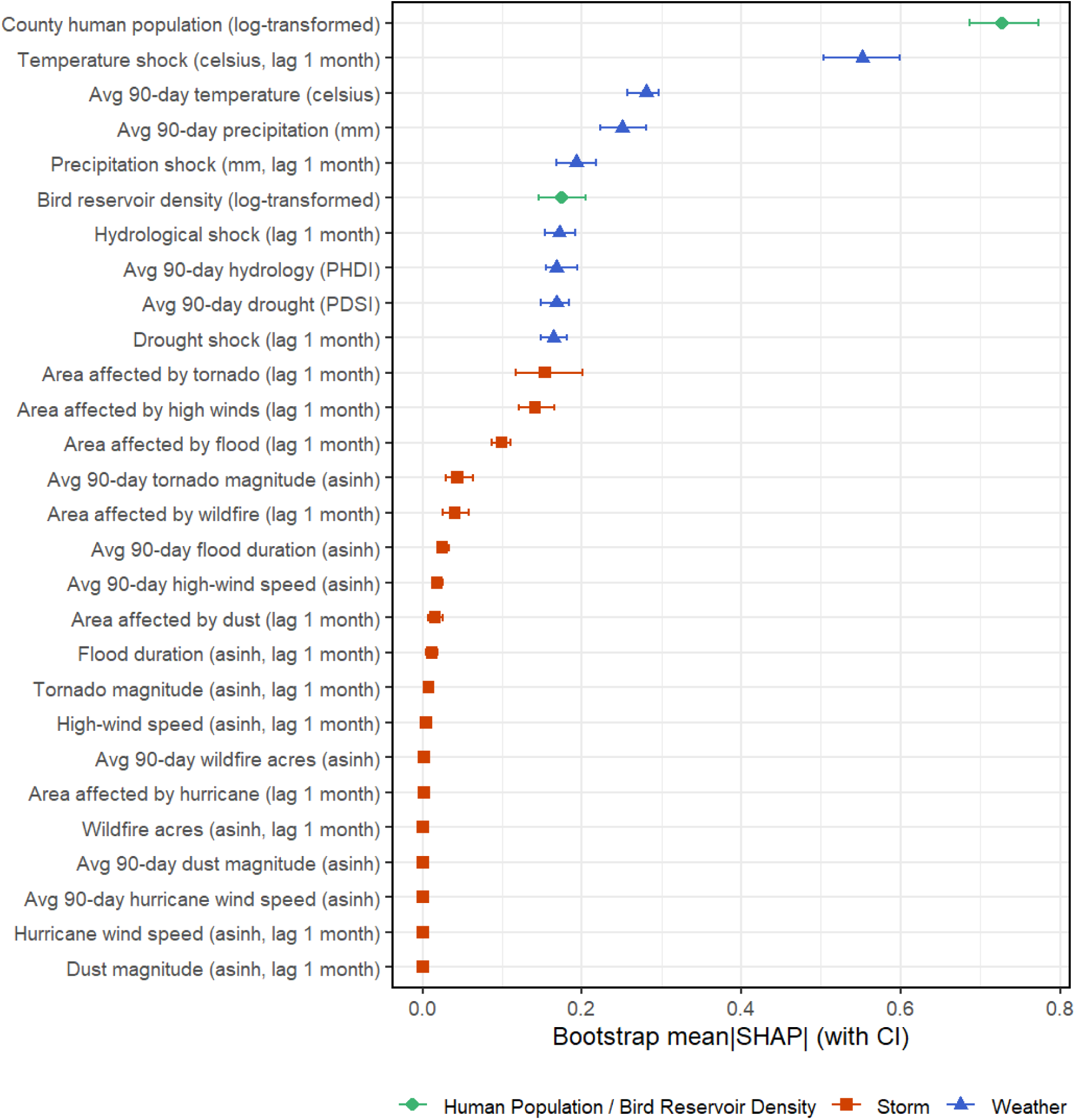
Variable contribution to the prediction of H5N1 outbreak risk in US counties between 2022 - 2024. Points are the absolute value of the mean contribution to predictions (mean SHAP value) for the covariate across all data points (i.e., global feature contribution). Bars represent the 95% confidence interval across 25 bootstrapping iterations.

PDPs for the top 12 performing predictors illustrate how predicted county-level H5N1 outbreak risk varied with human population, bird activity and with weather and storm conditions (Figure 3). Across PDPs, uncertainty bands were narrowest in the central covariate ranges and widened toward the extremes, indicating greater stability of predicted effects where there was more data.

**Figure 3.**
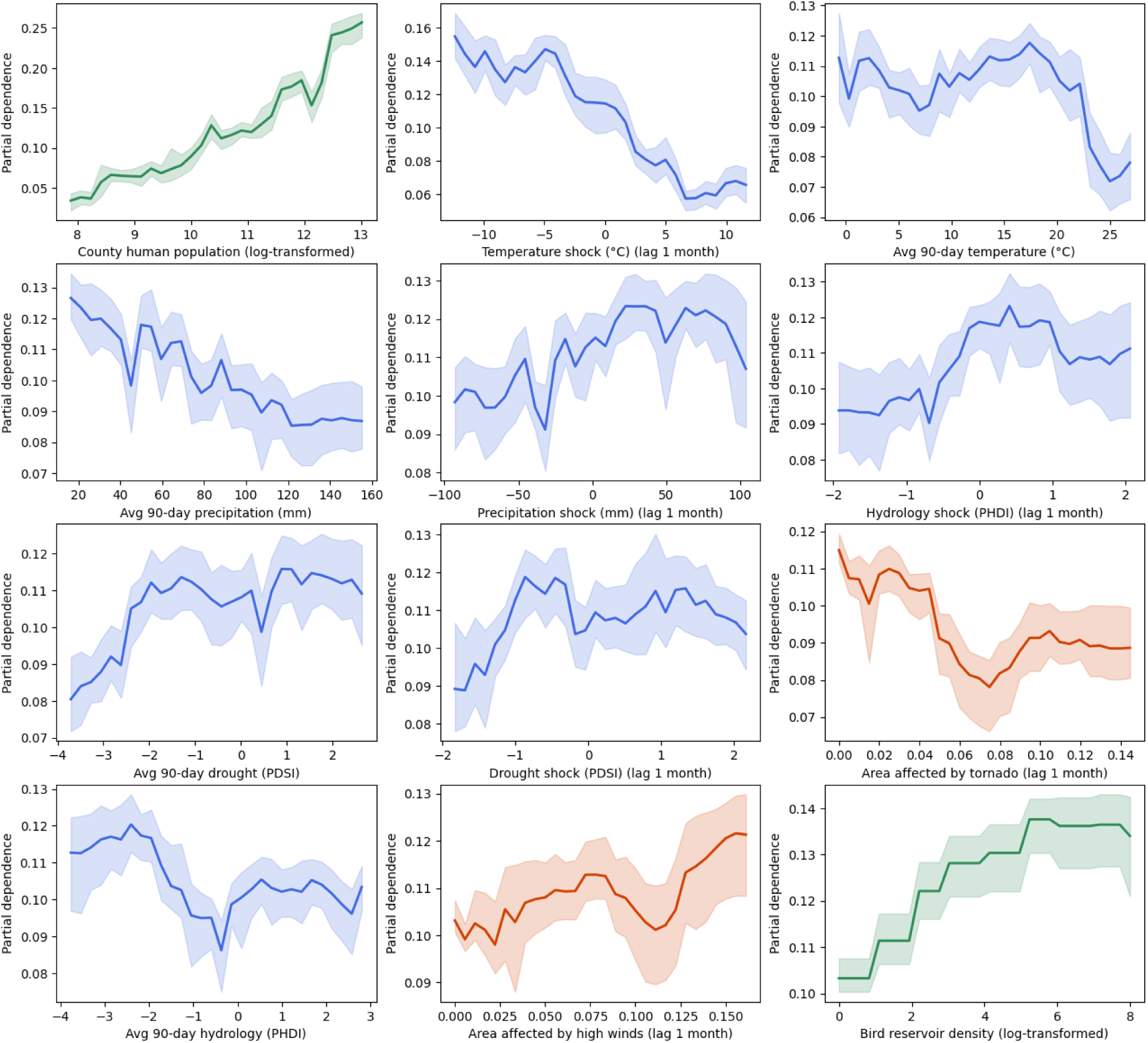
Partial dependence plots (PDPs) depicting the relationship between H5N1 outbreak risk and the top 12 predictors. For each univariate plot, blue lines represent the average probability of an outbreak after controlling all other covariates across 25 bootstrapped iterations. The y-axis scale differs between individual PDPs for visual clarity.

Risk increased monotonically with county-level human population, with a steeper rise at the upper end of the PDP distribution, and increased with bird reservoir density, showing rapid gains at low-to-moderate values followed by an apparent plateau at higher values. Warm temperature shocks were associated with lower outbreak risk while cold temperature shocks were associated with higher outbreak risk. The 90-day mean temperature also showed a similar trend, with lower temperatures generally associated with higher risk, dropping off steeply after a threshold value of around 18°C.

PDPs of the moisture-related variables (precipitation, drought, and hydrology) highlight more variability in their relationships with outbreak risk. For our three shock-related moisture variables, the driest conditions corresponded with periods of lower risk, while wetter conditions were generally found during periods of higher outbreak risk, though all plots featured relatively high variance, highlighted by jagged peaks and valleys and large uncertainty bands. A similar pattern was observed for 90-day mean drought, where risk was lowest in dry conditions and increased in wetter conditions. By contrast, 90-day mean precipitation and 90-day mean hydrology showed the opposite trend: outbreak risk was lowest during periods of wetter conditions, and highest during the driest periods.

Nearly all of our storm metrics were ranked lower than the included weather variables, with two exceptions: area affected by tornadoes and area affected by dust storms. The highest outbreak risk was found when tornado activity in surrounding counties was low, with risk declining sharply at moderate tornado activity levels, before rising slightly at the highest levels of surrounding tornado activity. In contrast, a higher area affected by high winds was associated with higher predicted outbreak risk, with risk increasing progressively as the area of high wind activity the month prior increased.

## 4. Discussion

Our study developed a high performing county-level predictive model of HPAI outbreak risk across the contiguous United States, integrating environmental, climatic, and anthropogenic predictors to characterize the drivers of H5N1 spread during the 2022–2024 panzootic. The model demonstrated strong discriminatory performance, and variable importance analyses consistently identified human population density and temperature-related variables as the most influential predictors of outbreak risk. The strong relationship that our model identified between human population and outbreak detection is likely multifactorial and warrants careful interpretation. Importantly, it does not wholly reflect poultry production intensity: commercial poultry operations in the United States are concentrated in predominantly rural areas, and the highest-population urban counties are generally not centers of poultry agriculture (51). Rather, the relationship may be more plausibly explained by detection and reporting dynamics: more populous counties tend to have greater access to veterinary infrastructure, diagnostic testing capacity, and surveillance resources that are necessary to identify and confirm HPAI cases. Additionally, virus establishment is influenced by the extent of surveillance and early detection, and therefore outbreak records are subject to an unknown degree of underreporting bias, which may be systematically lower in less-populated regions. Human population may also serve as a proxy for land use complexity: urbanization is characterized by rapid intensification of agriculture, socioeconomic change, and ecological fragmentation, which can have profound impacts on disease transmission (52). Our model indicated a steepening rise in outbreak risk at high population levels, further suggesting that risk-associated processes may scale nonlinearly in more populous counties where connectivity and intensity of human-wildlife-livestock interactions, along with surveillance capacity, can jointly amplify apparent outbreak probability. Disentangling true epidemiological risk from detection probability in the population signal remains an important direction for future work.

Temperature shock and 90-day average temperature were consistently ranked our second and third most influential predictors respectively, suggesting that predicted outbreak risk is associated with both temperature shocks and more prolonged deviations in temperature. Higher shocks and 90-day average temperatures showed a protective relationship with outbreak risk: higher temperatures in the prior month corresponded to lower predicted risk, while cold shocks were related with a rise in predicted risk. This pattern can be explained by several mechanisms. First, cooler prolonged conditions may enhance environmental persistence of influenza A viruses by prolonging viability in water, feces, and other environmental substrates, increasing opportunities for indirect transmission via shared water sources or contaminated fomites (53). Second, cold temperature shocks may alter wild bird host behavior and movements in ways that increase contact rates with domestic poultry (54). Thermoregulation is energetically expensive in wintering waterfowl, and persistent freezing temperatures cause wetlands to freeze, resulting in food and habitat limitations that displace waterfowl to more southerly locations (55). This displacement and concentration of birds into fewer viable habitats can increase contact rates with poultry operations. The timing and duration of migratory bird stopovers have been shown to influence the probability of HPAI infection in backyard poultry, further linking cold-driven behavioral changes to outbreak dynamics. Additionally, the energetic demands of cold-weather foraging may suppress immune function in wild birds: prolonged and intense exercise associated with migration has been shown to lead to immunosuppression, and migratory performance is negatively affected by infection, raising the possibility that cold-stressed birds may shed viruses at higher rates or for longer durations (56). Third, prolonged and abrupt cold conditions can change poultry management practices such as increased indoor confinement, reduced ventilation, greater crowding, and altered litter and moisture conditions, potentially increasing within-flock transmission and amplifying outbreaks once they begin. These management responses may also coincide with operational disruptions that affect biosecurity and detection efforts. Taken together, these mechanisms suggest that cold temperature shocks plausibly act through multiple pathways (environmental persistence, host behavior and management practices) to elevate outbreak risk, which may account in part for the strong predictive signal we observed.

Compared to the clear, directional patterns observed for temperature and population, moisture-related variables (precipitation, drought and hydrological conditions) showed more variables and, in some cases, discordant relationships with outbreak risk. Among all shock-based moisture variables and 90-day mean drought, wetter conditions generally correspond to higher predicted risk. This may be explained by surface water playing a well-established role in H5N1 transmission, as viral particles can be highly concentrated in fecal particles, which remain infective for extended periods when deposited in water supplies, allowing transmission to domestic livestock (35). Increased surface water availability from precipitation anomalies may therefore expand the environmental reservoir of viable viruses and increase the geographic overlap between wild waterfowl habitats and poultry operations.

In contrast, other 90-day mean moisture predictors (precipitation and hydrological conditions) showed an inverse pattern: this time prolonged dry conditions were associated with higher predicted outbreak risk. This apparent paradox may be explained by the behavioral ecology of water-scarce environments. Prolonged dry conditions increase the amount of energy needed for wild birds to survive and reproduce (57). Wild birds may be forced to travel greater ranges to forage for food, water and shelter. If there are fewer resources available than the wild bird requires, they may experience allostatic overload which in turn leads to suppression of immune function and increased vulnerability to viral infection and shedding (56). For H5N1 specifically, these adaptations may also increase interactions between reservoir hosts and other animals as not only do they increase their foraging range, but they may end up clustering around remaining water sources. When wild waterfowl concentrates at limited ponds or farm water sources during prolonged dry periods, the probability of contact with domestic poultry — and of fecal-oral transmission via shared or contaminated water — may increase.

Among storm-related variables, only those aggregated at the state rather than county level ranked among the top half of predictors. Of the state-level storm variables included, tornadoes and high winds were the most important and showed different relationships with outbreak risk, which may reflect distinct mechanisms. Higher predicted outbreak risk was associated with counties in a state experiencing a greater area of high-wind events, while the inverse held for tornadoes. One possible interpretation is that high-wind conditions, which include relatively modest sustained winds rather than the extreme conditions associated with tornadoes, may facilitate aerosol dispersal of contaminated fecal particulate matter between wild bird habitats, poultry operations, and their surrounding environments without triggering large-scale avian avoidance behavior. There is growing evidence that HPAI can be transmitted between poultry operations via wind-borne contaminated particles, and that mechanically ventilated buildings with high bird densities may facilitate windborne transmission even at viral concentrations below the minimum infectious dose (58–63). Fine dust particles of 2.5 μm or smaller carrying viral particles are capable of traveling hundreds of miles, suggesting that even moderate wind conditions could extend the effective transmission radius of contaminated material (64). By contrast, tornadoes are accompanied by severe drops in barometric pressure and infrasound, cues to which birds are demonstrably sensitive (65–67). Research has documented long-distance migrant birds undertaking facultative evacuation migrations more than 24 hours before the arrival of tornadic storms, with infrasound radiating over 1,000 km from tornadic systems identified as the likely sensory warning (68). Radar data also showed birds taking flight to avoid tornado formations (69). These triggers may effectively reduce the geographic overlap between migratory wild birds and domestic poultry in tornado-prone areas, attenuating rather than elevating spillover risk. Another mechanism for the overall decrease in risk is that tornadoes may also disrupt biosecurity, farm operations, or surveillance efforts. These contrasting patterns suggest that different categories of storm activity may have fundamentally different effects on the spatial distribution and behavior of wild birds and viral aerosol exposure, and that treating storm variables as a homogeneous category may obscure meaningful heterogeneity in their epidemiological effects.

Finally, bird reservoir density was also an important predictor. Bird reservoir density showed a broadly positive and monotonically increasing relationship with predicted outbreak risk, with the PDP revealing a steep stepwise rise from low to moderate density values that gradually plateaus at higher densities. This finding is consistent with the foundational ecology of HPAI spillover, wherein wild waterfowl, particularly Anseriformes, are the primary reservoir for avian influenza viruses and the principal source of wild-to-domestic introductions in the current North American panzootic. The mechanistic pathway is well established: waterfowl such as dabbling ducks exhibit high viral shedding rates, which can result in the accumulation of large viral loads in shared aquatic environments and perpetuate disease spread (70, 71). In addition, farm-level studies have identified the presence of waterfowl in surrounding areas as an important risk factor for HPAI infection in commercial poultry operations (72, 73). This finding reinforces that wild bird reservoirs are major drivers of H5N1 risk and suggests that spatiotemporally resolved waterfowl abundance data should remain a central input in future county-level risk surveillance frameworks.

Overall, our model achieved an AUC-PR of 0.237, representing a roughly 7-fold improvement over a naive random classifier, indicating that the spatiotemporal structure of HPAI outbreak risk is at least partially predictable from routinely available surveillance and environmental data. In practical terms, this suggests that the model has learned generalizable signals from the weather, storm, bird density, and population predictors included. Although our model’s precision of 0.181 means that approximately 1 in 6 county-months that were flagged by our model as high-risk would ultimately have an HPAI outbreak, our model’s high recall of 0.726 highlight’s the model’s ability to correctly flag approximately 73% of true outbreak county-months, a meaningful sensitivity for an early warning application where missing an outbreak carries substantial health risks and economic consequences. Practically, these results indicate that our model could serve as a high-recall early-warning screen to help focus surveillance resources and inform targeted biosecurity interventions; depending on response capacity, the alert threshold can be tuned to trade some sensitivity for higher precision in resource-constrained deployments or retained for maximal case-finding when missing outbreaks is costlier than investigating false alarms.

Our results should be interpreted with several important caveats. The most fundamental limitation of this work is that, while GBMs and PDPs can characterize complex, non-linear relationships between predictors and outcomes, they do not establish causality. Our analysis did not explicitly test causal pathways, and the relationships identified here may reflect correlational structure rather than underlying mechanisms. GBMs are additionally sensitive to sampling bias, class imbalance, and temporal leakage, all of which may inflate apparent relationships if not carefully addressed. Our dataset was substantially imbalanced (outbreak-positive observations were considerably rarer than outbreak-negative ones) and while we took many steps to address this, residual imbalance may have influenced model behavior and the interpretation of predictor importance. A related concern is that population density and some surveillance-linked covariates may partly encode detection and reporting intensity rather than true outbreak incidence. HPAI outbreaks in poultry, for instance, can only be detected in counties where poultry facilities exist and are subject to surveillance, meaning that apparent geographic variation in risk may partly reflect the spatial distribution of monitoring infrastructure rather than underlying transmission dynamics. Future analyses would benefit from incorporating explicit indicators of surveillance effort as covariates to partially disentangle true incidence from detection probability.

Further limitations lie in how the outcome variable was constructed. By aggregating wild bird detections with commercial poultry and backyard flocks into a single binary outcome, we implicitly treated these as manifestations of the same process with shared timing and mechanisms of infection. This assumption is likely violated in practice: wild bird detections and backyard flocks can precede commercial poultry outbreaks (74) and may serve as early-warning signals of spillover events rather than equivalent outcomes (75). Similarly, binarizing the outcome variable means that a county experiencing a single detection and one experiencing repeated outbreaks within the same time window are treated identically, obscuring variation in outbreak severity and burden. Future investigations should disaggregate outbreak types and consider ordinal or count-based outcome specifications to better capture heterogeneity in transmission dynamics and severity.

Finally, the rarity of severe weather events at the county level likely limited our ability to detect relationships between storm events and outbreak risk. Highlighting this point is the fact that only those storm events that were aggregated at the state, rather than county level, ranked among the top half of predictors in our model. We believe this likely reflects a statistical power limitation: severe storm events such as tornadoes are rare at the individual county level, and with only a few years of outbreak data available, county-level storm exposure may be too sparse to resolve a reliable signal. As HPAI surveillance data accumulate, revisiting storm predictors at finer spatial scales and across longer time horizons will be an important priority. More broadly, future work should explore alternative lag windows and spatial scales, including migratory bird flyway-level aggregations, and evaluate predictor interactions such as EWEs by bird reservoir presence, or population density by poultry density, to better characterize the potential mechanistic pathways highlighted in this work.

## 5. Conclusions

This investigation demonstrates that routinely available weather, storm, bird density and human population data can be integrated within a machine learning framework predict HPAI H5N1 outbreak risk at the county-month level across the United States. Our GBM classifier achieved strong discriminative performance, and partial dependence analyses revealed interpretable, biologically plausible relationships between outbreak risk and key predictors including population density, bird activity, temperature shocks, and moisture-related indices. The divergent effects of short-term versus prolonged moisture exposure, and the contrasting relationships of tornado and high wind activity with outbreak risk, highlight the importance of temporal scale and exposure specificity when characterizing environmental drivers of zoonotic spillover. Taken together, these findings suggest that spatially and temporally resolved environmental surveillance data could inform early warning systems, targeted biosecurity interventions, and the pre-positioning of response resources ahead of outbreak events. Future work should prioritize disaggregating outbreaks into wild birds, commercial poultry and backyard flock categories. In addition, future investigations should also account for surveillance effort, extend observation windows to better characterize rare weather exposures, and evaluate interaction effects between bird activity and environmental stressors. As HPAI continues to circulate in wild bird reservoirs, and periodic spillover into domestic poultry remains a persistent threat to agricultural systems and public health, predictive modeling approaches of this kind represent a scalable and actionable complement to traditional outbreak surveillance.

## CRediT author statement

**William W. Zou:** Conceptualization, Methodology, Software, Validation, Formal Analysis, Investigation, Data Curation, Writing – Original draft, Writing – Review & Editing, Visualization, Funding acquisition. **Elise N. Grover**: Conceptualization, Methodology, Investigation, Resources, Writing – Original draft, Writing – Review & Editing, Visualization, Supervision, Project Administration, Funding acquisition. **Elizabeth J. Carlton**: Resources, Writing - Review & Editing, Supervision, Project administration.

## Data Availability

The data that support the findings of this study were derived from the following resources, available in the public domain. Wild Bird HPAI Detection data are available from the US Department of Agriculture Animal and Plan Health Inspection Service (USDA APHIS) Detections of Highly Pathogenic Avian Influenza in Wild Birds database [Internet], available at: https://www.aphis.usda.gov/livestock-poultry-disease/avian/avian-influenza/hpai-detections/wild-birds?page=1 Commercial and backyard HPAI confirmation data are available from the US Department of Agriculture Animal and Plan Health Inspection Service (USDA APHIS) Confirmations of Highly Pathogenic Avian Influenza in Commercial and Backyard Flocks database [Internet], available at: https://www.aphis.usda.gov/livestock-poultry-disease/avian/avian-influenza/hpai-detections/commercial-backyard-flocks US monthly climate data summaries are available from the National Oceanic and Atmospheric Administration (NOAA) Monthly U.S. Climate Divisional Database (NClimDiv) [Internet], available at: https://www.ncei.noaa.gov/access/metadata/landing-page/bin/iso?id=gov.noaa.ncdc:C00005 doi:10.7289/V5M32STR US Storm event data are available from the National Oceanic and Atmospheric Administration (NOAA) Storm Event Database [Internet], available at: https://www.ncei.noaa.gov/stormevents/ftp.jsp Bird density data are available from the US Geological Service (USGS) Bird Banding Laboratory's Data Exploration Tool [Internet], available at: https://www.usgs.gov/labs/bird-banding-laboratory/data

## Abbreviations

EWEs: Extreme Weather Events
HPAI: Highly Pathogenic Avian Influenza
GBM: Gradient Boosting Machine
USDA: United States Department of Agriculture
APHIS: Animal and Plant Health Inspection Service
PDSI: Palmer Drought Severity Index
PHDI: Palmer Hydrological Drought Index
NOAA: National Oceanic Atmospheric Association
BBL: Bird Banding Lab
AUC-PR: Area Under the Precision-Recall Curve
ROC-AUC: Receiver Operating Characteristic Area Under the Curve
SHAP: SHapley Additive exPlanations
PDPs: Partial Dependence Plots
SD: Standard Deviation

## References

1. Meadows AJ, Stephenson N, Madhav NK, Oppenheim B. Historical trends demonstrate a pattern of increasingly frequent and severe spillover events of high-consequence zoonotic viruses. BMJ Global Health. 2023;8(11).

2. Baker RE, Mahmud AS, Miller IF, Rajeev M, Rasambainarivo F, Rice BL, et al. Infectious disease in an era of global change. Nature reviews microbiology. 2022;20(4):193–205.

3. Semenza JC, Rocklöv J, Ebi KL. Climate change and cascading risks from infectious disease. Infectious diseases and therapy. 2022;11(4):1371–90.

4. Seneviratne SI, X. Zhang, M. Adnan, W. Badi, C. Dereczynski, A. Di Luca, S. Ghosh, I. Iskandar, J. Kossin, S. Lewis, F. Otto, I. Pinto, M. Satoh, S.M. Vicente-Serrano, M. Wehner, and B. Zhou. Weather and Climate Extreme Events in a Changing Climate. In Climate Change 2021: The Physical Science Basis. Contribution of Working Group I to the Sixth Assessment Report of the Intergovernmental Panel on Climate Change. 2021.

5. Faust CL, Castellanos AA, Peel AJ, Eby P, Plowright RK, Han BA, et al. Environmental variation across multiple spatial scales and temporal lags influences Hendra virus spillover. Journal of Applied Ecology. 2023;60(7):1457–67.

6. Liao H, Lyon CJ, Ying B, Hu T. Climate change, its impact on emerging infectious diseases and new technologies to combat the challenge. Emerging microbes & infections. 2024;13(1):2356143.

7. Wang Z, Pei S, Cui H, Zhang J, Jia Z. Zoonotic spillover and extreme weather events drive the global outbreaks of airborne viral emerging infectious diseases. Journal of Medical Virology. 2024;96(6):e29737.

8. Ison MG, Marrazzo J. The emerging threat of H5N1 to human health. Mass Medical Soc; 2025. p. 916–8.

9. Clark L, Hall J. Avian influenza in wild birds: status as reservoirs, and risks to humans and agriculture. Ornithological monographs. 2006:3–29.

10. Couty M, Guinat C, Fornasiero D, Briand F-X, Henry P-Y, Grasland B, et al. The role of wild birds in the global highly pathogenic avian influenza H5 panzootic, 2020–2023. npj Biodiversity. 2026;5(1):1.

11. Lycett SJ, Duchatel F, Digard P. A brief history of bird flu. Philosophical Transactions of the Royal Society B: Biological Sciences. 2019;374(1775).

12. Charostad J, Rukerd MRZ, Mahmoudvand S, Bashash D, Hashemi SMA, Nakhaie M, et al. A comprehensive review of highly pathogenic avian influenza (HPAI) H5N1: An imminent threat at doorstep. Travel medicine and infectious disease. 2023;55:102638.

13. Krammer F, Hermann E, Rasmussen AL. Highly pathogenic avian influenza H5N1: history, current situation, and outlook. Journal of Virology. 2025;99(4):e02209–24.

14. Van Kerkhove MD, Mumford E, Mounts AW, Bresee J, Ly S, Bridges CB, et al. Highly pathogenic avian influenza (H5N1): pathways of exposure at the animal human interface, a systematic review. PloS one. 2011;6(1):e14582.

15. Saenz HSC, Cruz-Ausejo L. Preventive, safety and control measures against Avian Influenza A (H5N1) in occupationally exposed groups: A scoping review. One Health. 2024;19:100766.

16. Avian Influenza Weekly Update # 1037: 13 March 2026. World Health Organization Western Pacific Regional Emergencies Programme and Division of Health Security and Emergencies; 2025.

17. Peacock TP, Moncla L, Dudas G, VanInsberghe D, Sukhova K, Lloyd-Smith JO, et al. The global H5N1 influenza panzootic in mammals. Nature. 2025;637(8045):304–13.

18. Caliendo V, Lewis NS, Pohlmann A, Baillie SR, Banyard AC, Beer M, et al. Transatlantic spread of highly pathogenic avian influenza H5N1 by wild birds from Europe to North America in 2021. Scientific reports. 2022;12(1):11729.

19. Center NWH. Avian Influenza Surveillance: United States Geological Service; 2025 [Available from: https://www.usgs.gov/centers/nwhc/science/avian-influenza-surveillance?

20. Avian Influenza A (H5N5) - United States of America: World Health Organization; 2025 [Available from: https://www.who.int/emergencies/disease-outbreak-news/item/2025-DON590.

21. First H5 Bird Flu Death Reported in United States [press release]. CDC Newsroom: US Center for Disease Control2025.

22. Mena A, von Fricken ME, Anderson BD. The impact of Highly Pathogenic Avian Influenza H5N1 in the United States: A scoping review of past detections and present outbreaks. Viruses. 2025;17(3):307.

23. Garg S, Reinhart K, Couture A, Kniss K, Davis CT, Kirby MK, et al. Highly pathogenic avian influenza A (H5N1) virus infections in humans. New England Journal of Medicine. 2025;392(9):843–54.

24. Signore AV, Giacinti J, Jones ME, Erdelyan CN, McLaughlin A, Alkie TN, et al. Spatiotemporal reconstruction of the North American A (H5N1) outbreak reveals successive lineage replacements by descendant reassortants. Science Advances. 2025;11(28):eadu4909.

25. Kandeil A, Patton C, Jones JC, Jeevan T, Harrington WN, Trifkovic S, et al. Rapid evolution of A (H5N1) influenza viruses after intercontinental spread to North America. Nature communications. 2023;14(1):3082.

26. Ashley E, Vanstreels RET, Barbieri M, Puryear W, Gulland F, Field C, et al. High pathogenicity avian influenza in pinniped conservation. Philosophical Transactions of the Royal Society B: Biological Sciences. 2026;381(1945).

27. Fabrizio TP, Kandeil A, Harrington WN, Jones JC, Jeevan T, Andreev K, et al. Genotype B3. 13 influenza A (H5N1) viruses isolated from dairy cattle demonstrate high virulence in laboratory models, but retain avian virus-like properties. Nature Communications. 2025;16(1):6771.

28. Zhang X, Lam SJ-A, Chen L-L, Fong CH-Y, Chan W-M, Ip JD, et al. Avian influenza virus A (H5N1) genotype D1. 1 is better adapted to human nasal and airway organoids than genotype B3. 13. The Journal of Infectious Diseases. 2026;233(3):e662–e6.

29. Chrzastek K, Lieber CM, Plemper RK. H5n1 clade 2.3. 4.4 B: evolution, global spread, and host range expansion. Pathogens. 2025;14(9):929.

30. Bhojani MS, Bhojani MF. The emerging pandemic threat of H5N1: Evolutionary adaptations for human transmission, zoonotic spillovers and surveillance gaps. Gene. 2025:149723.

31. Prosser DJ, Teitelbaum CS, Yin S, Hill NJ, Xiao X. Climate change impacts on bird migration and highly pathogenic avian influenza. Nature Microbiology. 2023;8(12):2223–5.

32. Tietze DT. Bird species: how they arise, modify and vanish: Springer; 2018.

33. Ndachena N, Toorians ME, Farrell MJ, Govender D, Davies TJ. Effect of drought on wildlife activity at artificial waterholes. Biological Conservation. 2025;310:111370.

34. Puig-Gironès R, Brotons L, Pons P, Franch M. Examining the temporal effects of wildfires on forest birds: Should I stay or should I go? Forest Ecology and Management. 2023;549:121439.

35. Martin G, Becker DJ, Plowright RK. Environmental persistence of influenza H5N1 is driven by temperature and salinity: insights from a Bayesian meta-analysis. Frontiers in Ecology and Evolution. 2018;6:131.

36. Ahrens AK, Selinka H-C, Wylezich C, Wonnemann H, Sindt O, Hellmer HH, et al. Investigating environmental matrices for use in avian influenza virus surveillance—surface water, sediments, and avian fecal samples. Microbiology spectrum. 2023;11(2):e02664–22.

37. Nazir J, Haumacher R, Ike AC, Marschang RE. Persistence of avian influenza viruses in lake sediment, duck feces, and duck meat. Applied and environmental microbiology. 2011;77(14):4981–5.

38. Confirmations of Highly Pathogenic Avian Influenza in Commercial and Backyard Flocks: Animal and Plan Health Inspection Service US Department of Agriculture; 2022 [Available from: https://www.aphis.usda.gov/livestock-poultry-disease/avian/avian-influenza/hpai-detections/commercial-backyard-flocks.

39. Climate at a Glance County Mapping. In: Administration NOaA, editor. National Centers for Environmental Information1895–2026.

40. Vose RSA, Scott; Squires, Mike; Durre, Imke; Menne, Matthew J.; Williams, Claude N., Jr.; Fenimore, Chris; Gleason, Karin; Arndt, Derek. NOAA Monthly U.S. Climate Divisional Database (NClimDiv). In: Center NNCD, editor. National Centers for Environmental Information: National Oceanic and Atmospheric Administration; 2014.

41. Storm Events Database. In: Administration NOaA, editor. National Centers for Environmental Information.

42. Smith GJ. The US Geological Survey Bird Banding Laboratory: An integrated scientific program supporting research and conservation of North American birds. US Geological Surey; 2013. Report No.: 2331-1258.

43. Avian Influenza: Affected Bird Species. Library & Archives: Memorial Sloan Kettering Cancer Center.

44. UC Census Bureau Releases 2018-2022 ACS 5-Year Estimates. In: Bureau UC, editor.: US Census Bureau; 2023.

45. Isaac NJ, Jarzyna MA, Keil P, Dambly LI, Boersch-Supan PH, Browning E, et al. Data integration for large-scale models of species distributions. Trends in ecology & evolution. 2020;35(1):56–67.

46. Konstantinov AV, Utkin LV. Interpretable machine learning with an ensemble of gradient boosting machines. Knowledge-Based Systems. 2021;222:106993.

47. Chen T, Guestrin C, editors. Xgboost: A scalable tree boosting system. Proceedings of the 22nd acm sigkdd international conference on knowledge discovery and data mining; 2016.

48. Saito T, Rehmsmeier M. The precision-recall plot is more informative than the ROC plot when evaluating binary classifiers on imbalanced datasets. PloS one. 2015;10(3):e0118432.

49. Molnar C. Interpretable Machine Learning: A Guide for Making Black Box Models Explainable. Third Edition ed2025.

50. Ergün S. Explaining xgboost predictions with shap value: a comprehensive guide to interpreting decision tree-based models. New Trends in Computer Sciences. 2023;1(1):19–31.

51. Bruhn MC, Munoz B, Cajka J, Smith G, Curry RJ, Wagener DK, et al. Synthesized population databases: a geospatial database of US poultry farms. Methods report (RTI Press). 2012:1.

52. Hassell JM, Begon M, Ward MJ, Fèvre EM. Urbanization and disease emergence: dynamics at the wildlife–livestock–human interface. Trends in ecology & evolution. 2017;32(1):55–67.

53. Paek M, Lee Y, Yoon H, Kang H, Kim M, Choi J, et al. Survival rate of H5N1 highly pathogenic avian influenza viruses at different temperatures. Poultry science. 2010;89(8):1647–50.

54. Reperant LA, Fučkar NS, Osterhaus AD, Dobson AP, Kuiken T. Spatial and temporal association of outbreaks of H5N1 influenza virus infection in wild birds with the 0 C isotherm. PLoS pathogens. 2010;6(4):e1000854.

55. Xu F, Si Y. The frost wave hypothesis: How the environment drives autumn departure of migratory waterfowl. Ecological Indicators. 2019;101:1018–25.

56. Cheville N. Environmental factors affecting the immune response of birds: A review. Avian Diseases. 1979;23(2):308–14.

57. Wingfield JC, Pérez JH, Krause JS, Word KR, González-Gómez PL, Lisovski S, et al. How birds cope physiologically and behaviourally with extreme climatic events. Philosophical Transactions of the Royal Society B: Biological Sciences. 2017;372(1723).

58. Guinat C, Rouchy N, Camy F, Guérin JL, Paul MC. Exploring the wind-borne spread of highly pathogenic avian influenza H5N8 during the 2016–2017 epizootic in France. Avian Diseases. 2019;63(1s):246–8.

59. de Vos CJ, Elbers AR. Quantitative risk assessment of wind-supported transmission of highly pathogenic avian influenza virus to Dutch poultry farms via fecal particles from infected wild birds in the environment. Pathogens. 2024;13(7):571.

60. Ssematimba A, Hagenaars TJ, de Jong MC. Modelling the wind-borne spread of highly pathogenic avian influenza virus between farms. PLoS One. 2012;7(2):e31114.

61. Elbers AR, Gonzales JL, Koene MG, Germeraad EA, Hakze-van der Honing RW, van der Most M, et al. Monitoring wind-borne particle matter entering poultry farms via the air-inlet: Highly pathogenic avian influenza virus and other pathogens risk. Pathogens. 2022;11(12):1534.

62. Jonges M, Van Leuken J, Wouters I, Koch G, Meijer A, Koopmans M. Wind-mediated spread of low-pathogenic avian influenza virus into the environment during outbreaks at commercial poultry farms. PloS one. 2015;10(5):e0125401.

63. Nagy A, Černíková L, Sedlák K. Genetic data and meteorological conditions suggesting windborne transmission of H5N1 high-pathogenicity avian influenza between commercial poultry outbreaks. Plos one. 2025;20(9):e0319880.

64. Nguyen X, Zhao Y, Lin J, Purswell J, Tabler T, Voy B, et al. Modeling long-distance airborne transmission of highly pathogenic avian influenza carried by dust particles. Scientific Reports. 2023;13(1):16255.

65. Patrick SC, Assink JD, Basille M, Clusella-Trullas S, Clay TA, den Ouden OF, et al. Infrasound as a cue for seabird navigation. Frontiers in Ecology and Evolution. 2021;9:740027.

66. Zeyl JN, den Ouden O, Köppl C, Assink J, Christensen Dalsgaard J, Patrick SC, et al. Infrasonic hearing in birds: a review of audiometry and hypothesized structure–function relationships. Biological Reviews. 2020;95(4):1036–54.

67. Breuner CW, Sprague RS, Patterson SH, Woods HA. Environment, behavior and physiology: do birds use barometric pressure to predict storms? Journal of Experimental Biology. 2013;216(11):1982–90.

68. Streby HM, Kramer GR, Peterson SM, Lehman JA, Buehler DA, Andersen DE. Tornadic storm avoidance behavior in breeding songbirds. Current Biology. 2015;25(1):98–102.

69. Chilson PB, Daniel A, Cocks SB, Berkowitz DS, Melnikov V, Frick WF, et al., editors. The response of birds to abrupt natural hazards as observed using weather radar2012: ERAD.

70. Soda K, Tomioka Y, Usui T, Ozaki H, Ito H, Nagai Y, et al. Susceptibility of common dabbling and diving duck species to clade 2.3. 2.1 H5N1 high pathogenicity avian influenza virus: an experimental infection study. Journal of Veterinary Medical Science. 2023;85(9):942–9.

71. Papp Z, Clark RG, Parmley EJ, Leighton FA, Waldner C, Soos C. The ecology of avian influenza viruses in wild dabbling ducks (Anas spp.) in Canada. PloS one. 2017;12(5):e0176297.

72. Verhagen JH, Fouchier RA, Lewis N. Highly pathogenic avian influenza viruses at the wild–domestic bird interface in Europe: future directions for research and surveillance. Viruses. 2021;13(2):212.

73. King J, Staubach C, Lüder C, Koethe S, Günther A, Stacker L, et al. Connect to protect: dynamics and genetic connections of highly pathogenic avian influenza outbreaks in poultry from 2016 to 2021 in Germany. Viruses. 2022;14(9):1849.

74. Damodaran L, Jaeger AS, Moncla LH. Ecology and spread of the North American H5N1 epizootic. Nature. 2026 Jan 8;649(8096):432–41.

75. Global Consortium for H5N8 and Related Influenza Viruses. Role for migratory wild birds in the global spread of avian influenza H5N8. Science. 2016 Oct 14;354(6309):213–7.

