## Supplementary Materials for "Predicting highly pathogenic avian influenza H5N1 outbreak risk using extreme weather and bird migration data in machine learning models"

**Supplementary Table S1.** Predictor variables included in the gradient boosted machine model predicting H5N1 poultry outbreaks, their hypothesized direction of association, proposed mechanistic pathway, and supporting literature.

| **Predictor** | **Hypothesized Direction** | **Proposed Mechanism** |
| --- | --- | --- |
| **Avg 90-day temperature (°C)** | ↑ Risk (cold); ↓ Risk (warm) | Cold temperatures prolong environmental persistence of AIV in water, feces, and fomites by slowing viral degradation, extending the window of indirect transmission via shared water sources or contaminated surfaces. Sustained cold also imposes chronic thermoregulatory demands on wintering waterfowl, increasing allostatic load and potentially suppressing immune function, which may elevate viral replication and shedding rates. Prolonged freezing conditions additionally cause wetland habitats to ice over, concentrating birds into fewer open-water sources and increasing contact rates among wild birds and between wild birds and poultry operations. |
| **Temperature shock (°C) (lag 1 month)** | ↑ Risk (cold shock); ↓ Risk (hot shock) | Sudden cold anomalies impose acute physiological stress on migratory and resident waterfowl, triggering allostatic overload that can suppress immune defenses and amplify viral replication and shedding. Abrupt temperature drops may also trigger facultative southward movements, rapidly redistributing potentially infected birds into new geographic areas and increasing contact with naive poultry flocks. Cold shocks that cause sudden wetland freezing further compress available habitat, forcing birds to aggregate in higher densities around remaining open water, elevating transmission risk among wild birds and spillover risk to poultry. |
| **Avg 90-day precipitation (mm)** | ↑ Risk | Sustained wet conditions expand the availability and connectivity of wetland habitats, supporting longer and more frequent waterfowl stopovers and increasing the probability of contact between wild birds and poultry operations situated near water. Persistent surface water also extends the environmental viability of the H5N1 virus by maintaining the cool, aqueous conditions in which virus particles remain infectious, effectively broadening the environmental reservoir. Over longer periods, elevated moisture may also increase vegetation density around farms, providing cover that facilitates incursion of wild birds into poultry areas. |
| **Precipitation shock (mm) (lag 1 month)** | ↑ Risk | Acute precipitation events can rapidly create water bodies that attract aggregations of waterfowl, concentrating birds in areas that may not have previously posed spillover risk and increasing contact rates with nearby poultry. Flooding associated with heavy precipitation can breach farm biosecurity infrastructure — including fencing, drainage systems, and building integrity — and carry virus-laden water or fecal material into poultry facilities. Precipitation shocks may also displace birds from preferred foraging areas, forcing them into closer proximity with agricultural landscapes and intensifying human-wildlife-livestock interfaces. |
| **Hydrology shock (PHDI) (lag 1 month)** | ↑ Risk | Short-term water surplus conditions increase surface water availability across the landscape, expanding potential stopover and foraging habitat for migratory waterfowl and raising the probability of wild bird aggregation near poultry operations. Elevated surface water connectivity also facilitates waterborne dispersal of viral shed by wild birds, extending the effective transmission radius beyond direct contact. Sudden hydrological changes may additionally disrupt established bird movement patterns, redirecting flocks through areas of higher poultry density. |
| **Avg 90-day hydrology (PHDI)** | ↓ Risk | Prolonged water surplus conditions may paradoxically reduce outbreak risk by dispersing waterfowl across a wider and more heterogeneous landscape, reducing the density of birds at any given location and lowering per-encounter transmission probability. When high-quality stopover habitat is broadly available, birds are less compelled to aggregate at limited sites near agricultural areas, reducing wild bird–poultry contact rates. Additionally, sustained wet conditions may support healthier, less physiologically stressed bird populations with stronger immune function, potentially reducing individual shedding rates and viral prevalence in reservoir populations. |
| **Drought shock (PDSI) (lag 1 month)** | ↑ Risk (via bird deflection); ↑ Risk (via dust/arid surface) | Acute drought conditions reduce the availability of surface water and foraging habitat, forcing migratory and resident waterfowl to concentrate around remaining water sources — which may include farm ponds, irrigation infrastructure, and other agricultural water bodies — dramatically increasing contact rates with poultry. This behavioral compression also elevates allostatic load in affected birds: the energetic costs of extended foraging under resource-scarce conditions can trigger immune suppression, potentially increasing viral replication and shedding. Simultaneously, drought-induced arid surface exposure increases dust generation, which may mechanically transport virus-laden fecal particulates across farm boundaries and into poultry housing. |
| **Avg 90-day drought (PDSI)** | ↑ Risk | Prolonged drought conditions may reduce outbreak risk primarily through habitat displacement: sustained water scarcity pushes waterfowl away from traditionally used wetlands and migratory corridors, potentially redirecting birds away from poultry-dense agricultural regions rather than toward them. Over extended periods, severe drought may also reduce overall waterfowl population density in affected areas through reduced breeding success and habitat abandonment, lowering the regional reservoir burden. However, this apparent protective effect may reflect displacement rather than true risk reduction, as deflected birds may elevate risk elsewhere along the flyway. |
| **Wildfires**  **(lag 1 month and area affected)** | ↓ Risk | Wildfires destroy and degrade stopover habitat critical to migratory waterfowl, forcing birds to bypass affected areas and altering established flyway routes in ways that may reduce overlap with poultry-dense regions. The energetic cost of navigating around fire-affected landscapes and locating alternative stopover sites increases allostatic load in migrants, but the net effect on outbreak risk appears protective if birds are deflected away from agricultural areas entirely. Smoke and habitat loss may also temporarily suppress local wild bird activity and residency, reducing the density of potential reservoir hosts in fire-affected counties during and immediately following wildfire events. |
| **Flood hours (lag 1 month and area affected)** | ↑ Risk | Flooding can physically breach farm biosecurity infrastructure, including containment fencing, drainage channels, and building foundations, creating direct pathways for virus-laden floodwater to enter poultry facilities. Inundation of low-lying areas forces both wild birds and poultry into shared elevated terrain, increasing interspecies contact at a time when environmental contamination is high. Floodwaters also facilitate wide dispersal of viral shed by wild birds across the landscape, depositing infectious material in areas that may not otherwise serve as transmission hotspots. Recovery periods following flooding may further compromise biosecurity and surveillance capacity as farm operations are disrupted. |
| **Dust magnitude (lag 1 month and area affected)** | ↑ Risk | Dust storms may mechanically transport virus-laden particles across farm boundaries; associated with immunosuppressive physiological stress in wild birds during migration |
| **Hurricane wind speed (lag 1 month and area affected)** | ↑ Risk (acute disruption) | Hurricanes may physically destroy or severely damage poultry housing, breaching containment and exposing flocks to the environment at precisely the moment when viral risk from displaced wild birds may be elevated. Storm surge and associated flooding disperse virus-laden material across wide areas while simultaneously concentrating displaced wild birds and waterfowl in novel locations, increasing the geographic unpredictability of spillover risk. The severe energetic demands of surviving and navigating hurricane-force conditions impose acute allostatic overload on wild birds, potentially amplifying viral shedding in the post-storm period. Prolonged disruption to farm operations, veterinary services, and surveillance infrastructure in affected regions may further delay detection and containment of outbreaks that follow hurricane events. |
| **High wind speed**  **(lag 1 month and area affected)** | ↑ Risk | High wind conditions, distinct from the extreme and destructive forces of tornadoes, may facilitate the lofting and long-range dispersal of fine virus-laden particulate matter — including dried fecal dust and contaminated material — from wild bird aggregation sites and infected farms into neighboring poultry facilities. Mechanically ventilated high-density poultry buildings are particularly vulnerable, as intake airflow can draw contaminated aerosols directly into housing even at viral concentrations below conventional infectious dose thresholds. High winds may also increase the aerosolization of surface dust from arid or recently dried areas, amplifying mechanical transport of environmental viral loads. Unlike tornado-scale events, high winds are unlikely to trigger strong avoidance behavior in wild birds, meaning that elevated aerosol transmission risk is not offset by behavioral displacement of reservoir hosts. |
| **Tornado magnitude (lag 1 month and area affected)** | ↓ Risk | Contrary to an initial expectation of elevated risk, tornadoes may appear to reduce outbreak probability, likely through strong avoidance behavior in wild birds: research has documented that long-distance migrants initiate facultative evacuation movements more than 24 hours before tornadic storm arrival, responding to infrasound cues that radiate over 1,000 km from developing storm systems. This behavioral early departure reduces the geographic overlap between migratory waterfowl and poultry operations in tornado-affected areas at the critical moment of storm passage. Tornadoes may also disrupt farm operations and surveillance infrastructure, though this effect appears secondary to the dominant bird displacement mechanism. The net result suggests that tornado-associated wild bird avoidance behavior, rather than structural farm damage, may be the primary driver of the inverse risk relationship observed. |

**Supplementary Methods S1.** Rule Based String Extraction for Storm Events Narratives

“Strong wind” and “high wind” features from the NOAA database were combined into a single feature (windspeed > 20 knots), with their severity represented by their recorded maximum windspeed in knots. For all other storm features, we used string extraction to retrieve details on the magnitude of the event from a written narrative variable included for each event in the database. For hurricanes, we parsed narratives for wind-speed expressions (e.g., “sustained winds of … miles per hour (mph)”, “gusts up to … mph”) and set magnitude to the minimum mph mentioned across either narrative; when no numeric mph was present but a Saffir–Simpson category appeared, we input the category’s lower-bound wind speed (Category 1–5 : 74, 96, 111, 130, 157 mph respectively). All hurricane event narrative reports included either a numeric mph or a Saffir-Simpson category. For floods and dust storms, severity was calculated as the event duration in hours, using timestamps recording event start and end times because most flood and dust storm narrative reports did not include consistent metrics of severity such as flood height and volume, windspeed or area of impact. We recorded instances of non-positive flood and dust storm durations as missing, while flood and dust storm events that lasted less than an hour was recoded as one. For tornadoes, magnitude was computed as the tornado length multiplied by the tornado width, as many tornado narratives did not include windspeed. For wildfires, we extracted affected acreage mentions (e.g. “1,000 acres”) and used the lowest acreage value referenced.

**Supplementary Table S2.** Bird species susceptible to or documented as hosts of avian influenza A viruses, organized by epidemiological role that were incorporated into the *bird reservoir density* predictor. Domesticated poultry and captive wild birds represent direct agricultural exposure risks. Wild waterfowl (Anseriformes) serve as the primary natural reservoir, capable of maintaining and shedding viruses with minimal clinical signs. Accidental hosts include a broad range of wild birds that may acquire infection through environmental or direct contact. Scavengers and birds of prey are susceptible primarily through consumption of infected prey or carcasses. Scientific nomenclature follows current taxonomic conventions. Only bird species that have been observed in North America were included in this list. Source: Centers for Disease Control and Prevention (CDC).

| **Category** | **Common Name** | **Scientific Name** |
| --- | --- | --- |
| **Domesticated Poultry** | | |
| Domesticated Poultry | Duck | *Anas platyrhynchos domesticus* |
| Domesticated Poultry | Goose | *Anserinae sp.* |
| Domesticated Poultry | Chicken | *Gallus gallus domesticus* |
| Domesticated Poultry | Turkey | *Meleagris gallopavo* |
| **Captive Wild Birds** | | |
| Captive Wild Birds | Australian Bushturkey | *Alectura lathami* |
| Captive Wild Birds | Catalina Macaw | *Ara ararauna × Ara macao* |
| Captive Wild Birds | Scarlet Macaw | *Ara macao* |
| Captive Wild Birds | Military Macaw | *Ara militaris* |
| Captive Wild Birds | Common Quail | *Coturnix coturnix* |
| Captive Wild Birds | Burrowing Parrot | *Cyanoliseus patagonus* |
| Captive Wild Birds | Emu | *Dromaius novaeollandiae* |
| Captive Wild Birds | Blue Crane | *Grus paradisea* |
| Captive Wild Birds | Common Guineafowl | *Numida meleagris* |
| Captive Wild Birds | Peacock | *Pavo cristatus* |
| Captive Wild Birds | Grey Partridge | *Perdix perdix* |
| Captive Wild Birds | Common Pheasant | *Phasianus colchicus* |
| Captive Wild Birds | Chilean Flamingo | *Phoenicopterus chilensis* |
| Captive Wild Birds | Lesser Flamingo | *Phoenicopterus minor* |
| Captive Wild Birds | Ostrich | *Struthio camelus* |
| **Wild Reservoir & Host Species (Waterfowl)** | | |
| Wild Reservoir/Host | Mandarin Duck | *Aix galericulata* |
| Wild Reservoir/Host | Wood Duck | *Aix sponsa* |
| Wild Reservoir/Host | Egyptian Goose | *Alopochen aegyptiaca* |
| Wild Reservoir/Host | Brazilian Teal | *Amazonetta brasiliensis* |
| Wild Reservoir/Host | Northern Pintail | *Anas acuta* |
| Wild Reservoir/Host | American Green-winged Teal | *Anas carolinensis* |
| Wild Reservoir/Host | Northern Shoveler | *Anas clypeata* |
| Wild Reservoir/Host | Common Teal | *Anas crecca* |
| Wild Reservoir/Host | Blue-winged Teal | *Anas discors* |
| Wild Reservoir/Host | Falcated Duck | *Anas falcata* |
| Wild Reservoir/Host | Eurasian Wigeon | *Anas penelope* |
| Wild Reservoir/Host | Mallard | *Anas platyrhynchos* |
| Wild Reservoir/Host | Indian Spot-billed Duck | *Anas poecilorhyncha* |
| Wild Reservoir/Host | Garganey | *Anas querquedula* |
| Wild Reservoir/Host | American Black Duck | *Anas rubripes* |
| Wild Reservoir/Host | Gadwall | *Anas strepera* |
| Wild Reservoir/Host | Yellow-billed Duck | *Anas undulata* |
| Wild Reservoir/Host | Silver Teal | *Anas versicolor* |
| Wild Reservoir/Host | Greater White-fronted Goose | *Anser albifrons* |
| Wild Reservoir/Host | Greylag Goose | *Anser anser* |
| Wild Reservoir/Host | Pink-footed Goose | *Anser brachyrhynchus* |
| Wild Reservoir/Host | Snow Goose | *Anser caerulescens* |
| Wild Reservoir/Host | Lesser White-fronted Goose | *Anser erythropus* |
| Wild Reservoir/Host | Bean Goose | *Anser fabalis* |
| Wild Reservoir/Host | Ross's Goose | *Anser rossii* |
| Wild Reservoir/Host | Bean Goose | *Anser serrirostris* |
| Wild Reservoir/Host | Lesser Scaup | *Aythya affinis* |
| Wild Reservoir/Host | Redhead Duck | *Aythya americana* |
| Wild Reservoir/Host | Ring-necked Duck | *Aythya collaris* |
| Wild Reservoir/Host | Common Pochard | *Aythya ferina* |
| Wild Reservoir/Host | Tufted Duck | *Aythya fuligula* |
| Wild Reservoir/Host | Greater Scaup | *Aythya marila* |
| Wild Reservoir/Host | Ferruginous Pochard | *Aythya nyroca* |
| Wild Reservoir/Host | Wild Duck | *Aythyinae or Anatinae sp.* |
| Wild Reservoir/Host | Brent Goose | *Branta bernicla* |
| Wild Reservoir/Host | Canada Goose | *Branta canadensis* |
| Wild Reservoir/Host | Cackling Goose | *Branta hutchinsii* |
| Wild Reservoir/Host | Barnacle Goose | *Branta leucopsis* |
| Wild Reservoir/Host | Red-breasted Goose | *Branta ruficollis* |
| Wild Reservoir/Host | Nene | *Branta sandvicensis* |
| Wild Reservoir/Host | Common Goldeneye | *Bucephala clangula* |
| Wild Reservoir/Host | Muscovy Duck | *Cairina moschata* |
| Wild Reservoir/Host | Maned Duck | *Chenonetta jubata* |
| Wild Reservoir/Host | Coscoroba Swan | *Coscoroba coscoroba* |
| Wild Reservoir/Host | Black Swan | *Cygnus atratus* |
| Wild Reservoir/Host | Trumpeter Swan | *Cygnus buccinators* |
| Wild Reservoir/Host | Tundra Swan | *Cygnus columbianus* |
| Wild Reservoir/Host | Whooper Swan | *Cygnus cygnus* |
| Wild Reservoir/Host | Mute Swan | *Cygnus olor* |
| Wild Reservoir/Host | Plumed Whistling-Duck | *Dendrocygna eytoni* |
| Wild Reservoir/Host | Hooded Merganser | *Lophodytes cucullatus* |
| Wild Reservoir/Host | American Wigeon | *Mareca americana* |
| Wild Reservoir/Host | Marbled Teal | *Marmaronetta angustirostris* |
| Wild Reservoir/Host | Goosander | *Mergus merganser* |
| Wild Reservoir/Host | Red-crested Pochard | *Netta rufina* |
| Wild Reservoir/Host | Ruddy Duck | *Oxyura jamaicensis* |
| Wild Reservoir/Host | Spur-winged Goose | *Plectopterus gambensis* |
| Wild Reservoir/Host | Northern Shoveler | *Spatula clypeata* |
| Wild Reservoir/Host | Puna Teal | *Spatula puna* |
| Wild Reservoir/Host | Common Shelduck | *Tadorna tadorna* |
| **Accidental Hosts (Wading Birds, Shorebirds & Other Wild Birds)** | | |
| Accidental Host / Reservoir | Western Grebe | *Aechmophorus occidentalis* |
| Accidental Host / Reservoir | Red-winged Blackbird | *Agelaius phoeniceus* |
| Accidental Host / Reservoir | Razorbill | *Alca torda* |
| Accidental Host / Reservoir | Red-legged Partridge | *Alectoris rufa* |
| Accidental Host / Reservoir | Amazon Parrot | *Amazona farinose* |
| Accidental Host / Reservoir | Yellow-crowned Parrot | *Amazona ochrocephala* |
| Accidental Host / Reservoir | Sandhill Crane | *Antigone canadensis* |
| Accidental Host / Reservoir | White-naped Crane | *Antigone vipio* |
| Accidental Host / Reservoir | Green and Red Macaw | *Ara cloropterus* |
| Accidental Host / Reservoir | Great Egret | *Ardea alba* |
| Accidental Host / Reservoir | Grey Heron | *Ardea cinerea* |
| Accidental Host / Reservoir | Great Blue Heron | *Ardea herodias* |
| Accidental Host / Reservoir | Black-headed Heron | *Ardea melanocephala* |
| Accidental Host / Reservoir | Baer's Pochard | *Aythya baeri* |
| Accidental Host / Reservoir | Crowned Crane | *Balearica regulorum* |
| Accidental Host / Reservoir | Ruffed Grouse | *Bonasa umbellus* |
| Accidental Host / Reservoir | Eurasian Bittern | *Botaurus stellaris* |
| Accidental Host / Reservoir | Western Cattle Egret | *Bubulcus ibis* |
| Accidental Host / Reservoir | Barrow's Goldeneye | *Bucephala islandica* |
| Accidental Host / Reservoir | Green Heron | *Butorides virescens* |
| Accidental Host / Reservoir | Sanderling | *Calidris alba* |
| Accidental Host / Reservoir | Dunlin | *Calidris alpina* |
| Accidental Host / Reservoir | Red Knot | *Calidris canutus* |
| Accidental Host / Reservoir | Curlew Sandpiper | *Calidris ferruginea* |
| Accidental Host / Reservoir | Purple Sandpiper | *Calidris maritima* |
| Accidental Host / Reservoir | Little Stint | *Calidris minuta* |
| Accidental Host / Reservoir | Semipalmated Sandpiper | *Calidris pusilla* |
| Accidental Host / Reservoir | Black Guillemot | *Cepphus grille* |
| Accidental Host / Reservoir | Kentish Plover | *Charadrius alexandrines* |
| Accidental Host / Reservoir | Little Ringed Plover | *Charadrius dubius* |
| Accidental Host / Reservoir | Common Ringed Plover | *Charadrius hiaticula* |
| Accidental Host / Reservoir | Lesser Sand Plover | *Charadrius mongolus* |
| Accidental Host / Reservoir | Snowy Plover | *Charadrius nivosus* |
| Accidental Host / Reservoir | Chestnut-banded Plover | *Charadrius pallidus* |
| Accidental Host / Reservoir | Whiskered Tern | *Chlidonias hybrid* |
| Accidental Host / Reservoir | White-winged Black Tern | *Chlidonias leucoptera* |
| Accidental Host / Reservoir | Oriental Stork | *Ciconia boyciana* |
| Accidental Host / Reservoir | White Stork | *Ciconia ciconia* |
| Accidental Host / Reservoir | Stork | *Ciconiidae sp.* |
| Accidental Host / Reservoir | Eurasian Jackdaw | *Coloeus monedula* |
| Accidental Host / Reservoir | African Rock Pigeon | *Columba guinea* |
| Accidental Host / Reservoir | Rock Pigeon | *Columba livia* |
| Accidental Host / Reservoir | Common Wood-Pigeon | *Columba palumbus* |
| Accidental Host / Reservoir | Pigeon | *Columbidae sp.* |
| Accidental Host / Reservoir | Western Jackdaw | *Corvus monedula* |
| Accidental Host / Reservoir | Blue Jay | *Cyanocitta cristata* |
| Accidental Host / Reservoir | Montezuma Quail | *Cyrtonyx montezumae* |
| Accidental Host / Reservoir | Great Spotted Woodpecker | *Dendrocopos major* |
| Accidental Host / Reservoir | Little Egret | *Egretta garzetta* |
| Accidental Host / Reservoir | Snowy Egret | *Egretta thula* |
| Accidental Host / Reservoir | American White Ibis | *Eudocimus albus* |
| Accidental Host / Reservoir | Scarlet Ibis | *Eudocimus ruber* |
| Accidental Host / Reservoir | Common Chaffinch | *Fringilla coelebs* |
| Accidental Host / Reservoir | Atlantic Puffin | *Fratercula arctica* |
| Accidental Host / Reservoir | American Coot | *Fulica americana* |
| Accidental Host / Reservoir | Common Coot | *Fulica atra* |
| Accidental Host / Reservoir | Northern Fulmar | *Fulmarus glacialis* |
| Accidental Host / Reservoir | Common Moorhen | *Gallinula chloropus* |
| Accidental Host / Reservoir | Eurasian Jay | *Garrulus glandarius* |
| Accidental Host / Reservoir | Common Loon | *Gavia immer* |
| Accidental Host / Reservoir | Pacific Loon | *Gavia pacifica* |
| Accidental Host / Reservoir | Red-throated Loon | *Gavia stellata* |
| Accidental Host / Reservoir | Northern Bald Ibis | *Geronticus eremita* |
| Accidental Host / Reservoir | Common Crane | *Grus grus* |
| Accidental Host / Reservoir | Red-crowned Crane | *Grus japonensis* |
| Accidental Host / Reservoir | Hooded Crane | *Grus monacha* |
| Accidental Host / Reservoir | African Black Oystercatcher | *Haematopus moquini* |
| Accidental Host / Reservoir | Eurasian Oystercatcher | *Haematopus ostralegus* |
| Accidental Host / Reservoir | Black-winged Stilt | *Himantopus himantopus* |
| Accidental Host / Reservoir | Swallow | *Hirundo sp.* |
| Accidental Host / Reservoir | Little Gull | *Hydrocoloeus minutus* |
| Accidental Host / Reservoir | Caspian Tern | *Hydroprogne caspia* |
| Accidental Host / Reservoir | Dark-eyed Junco | *Junco hyemalis* |
| Accidental Host / Reservoir | Laughing Gull | *Leucophaeus atricilla* |
| Accidental Host / Reservoir | Munia | *Lonchura sp.* |
| Accidental Host / Reservoir | Smew | *Mergellus albellus* |
| Accidental Host / Reservoir | Scaly-sided Merganser | *Mergus squamatus* |
| Accidental Host / Reservoir | Pied Wagtail | *Motacilla alba* |
| Accidental Host / Reservoir | American Wood Stork | *Mycteria americana* |
| Accidental Host / Reservoir | Painted Stork | *Mycteria leucocephala* |
| Accidental Host / Reservoir | Double-crested Cormorant | *Nannopterum auritum* |
| Accidental Host / Reservoir | Neotropic Cormorant | *Nannopterum brasilianum* |
| Accidental Host / Reservoir | Eurasian Curlew | *Numenius arquata* |
| Accidental Host / Reservoir | Curlew | *Numenius sp.* |
| Accidental Host / Reservoir | Black-crowned Night-Heron | *Nycticorax nycticorax* |
| Accidental Host / Reservoir | House Sparrow | *Passer domesticus* |
| Accidental Host / Reservoir | Dalmatian Pelican | *Pelecanus crispus* |
| Accidental Host / Reservoir | American White Pelican | *Pelecanus erythrorhynchos* |
| Accidental Host / Reservoir | Brown Pelican | *Pelecanus occidentalis* |
| Accidental Host / Reservoir | Great White Pelican | *Pelecanus onocrotalus* |
| Accidental Host / Reservoir | Peruvian Pelican | *Pelecanus thagus* |
| Accidental Host / Reservoir | Double-crested Cormorant | *Phalacrocorax auritus* |
| Accidental Host / Reservoir | Cape Cormorant | *Phalacrocorax capensis* |
| Accidental Host / Reservoir | Great Cormorant | *Phalacrocorax carbo* |
| Accidental Host / Reservoir | Pygmy Cormorant | *Phalacrocorax pygmaeus* |
| Accidental Host / Reservoir | Common Pheasant | *Phasianus colchicus* |
| Accidental Host / Reservoir | Red-legged Cormorant | *Phalacrocorax gaimardi* |
| Accidental Host / Reservoir | Greater Flamingo | *Phoenicopterus roseus* |
| Accidental Host / Reservoir | Eurasian Spoonbill | *Platalea leucorodia* |
| Accidental Host / Reservoir | Black-faced Spoonbill | *Platalea minor* |
| Accidental Host / Reservoir | White-faced Ibis | *Plegadis chihi* |
| Accidental Host / Reservoir | Glossy Ibis | *Plegadis falcinellus* |
| Accidental Host / Reservoir | Southern Masked-Weaver | *Ploceus velatus* |
| Accidental Host / Reservoir | Horned Grebe | *Podiceps auritus* |
| Accidental Host / Reservoir | Great Crested Grebe | *Podiceps cristatus* |
| Accidental Host / Reservoir | Red-necked Grebe | *Podiceps grisegena* |
| Accidental Host / Reservoir | Eared Grebe | *Podiceps nigricollis* |
| Accidental Host / Reservoir | Great Shearwater | *Puffinus gravis* |
| Accidental Host / Reservoir | Manx Shearwater | *Puffinus puffinus* |
| Accidental Host / Reservoir | Boat-tailed Grackle | *Quiscalus major* |
| Accidental Host / Reservoir | Common Grackle | *Quiscalus quiscula* |
| Accidental Host / Reservoir | Water Rail | *Rallus aquaticus* |
| Accidental Host / Reservoir | Pied Avocet | *Recurvirostra avosetta* |
| Accidental Host / Reservoir | Greater Rhea | *Rhea americana* |
| Accidental Host / Reservoir | Black-legged Kittiwake | *Rissa tridactyla* |
| Accidental Host / Reservoir | Black Skimmer | *Rynchops niger* |
| Accidental Host / Reservoir | Eurasian Woodcock | *Scolopax rusticola* |
| Accidental Host / Reservoir | Common Eider | *Somateria mollissima* |
| Accidental Host / Reservoir | King Eider | *Somateria spectabilis* |
| Accidental Host / Reservoir | African Penguin | *Spheniscus demersus* |
| Accidental Host / Reservoir | Arctic Skua | *Stercorarius parasiticus* |
| Accidental Host / Reservoir | Great Skua | *Stercorarius skua* |
| Accidental Host / Reservoir | Roseate Tern | *Sterna dougallii* |
| Accidental Host / Reservoir | Forster's Tern | *Sterna forsteri* |
| Accidental Host / Reservoir | Common Tern | *Sterna hirundo* |
| Accidental Host / Reservoir | Arctic Tern | *Sterna paradisaea* |
| Accidental Host / Reservoir | Eurasian Collared Dove | *Streptopella decaocto* |
| Accidental Host / Reservoir | Laughing Dove | *Streptopella senegalensis* |
| Accidental Host / Reservoir | Common Starling | *Sturnus vulgaris* |
| Accidental Host / Reservoir | Little Grebe | *Tachybaptus ruficollis* |
| Accidental Host / Reservoir | Tree Swallow | *Tachycineta bicolor* |
| Accidental Host / Reservoir | Swift Tern | *Thalasseus bergii* |
| Accidental Host / Reservoir | Elegant Tern | *Thalasseus elegans* |
| Accidental Host / Reservoir | Royal Tern | *Thalasseus maximus* |
| Accidental Host / Reservoir | Sandwich Tern | *Thalasseus sandvicensis* |
| Accidental Host / Reservoir | Sacred Ibis | *Threskiornis aethiopicus* |
| Accidental Host / Reservoir | Wood Sandpiper | *Tringa glareola* |
| Accidental Host / Reservoir | Greater Yellowlegs | *Tringa melanoleuca* |
| Accidental Host / Reservoir | Green Sandpiper | *Tringa ochropus* |
| Accidental Host / Reservoir | Eurasian Blackbird | *Turdus merula* |
| Accidental Host / Reservoir | American Robin | *Turdus migratorius* |
| Accidental Host / Reservoir | Song Thrush | *Turdus philomelos* |
| Accidental Host / Reservoir | Fieldfare | *Turdus pilaris* |
| Accidental Host / Reservoir | Common Murre | *Uria aalge* |
| Accidental Host / Reservoir | Thick-billed Murre | *Uria lomvia* |
| Accidental Host / Reservoir | Northern Lapwing | *Vanellus vanellus* |
| **Scavengers & Birds of Prey** | | |
| Scavenger / Bird of Prey | Cooper's Hawk | *Accipiter cooperii* |
| Scavenger / Bird of Prey | Northern Goshawk | *Accipiter gentilis* |
| Scavenger / Bird of Prey | Eurasian Sparrowhawk | *Accipiter nisus* |
| Scavenger / Bird of Prey | Sharp-shinned Hawk | *Accipiter striatus* |
| Scavenger / Bird of Prey | Northern Saw-whet Owl | *Aegolius acadicus* |
| Scavenger / Bird of Prey | Golden Eagle | *Aquila chrysaetos* |
| Scavenger / Bird of Prey | Short-eared Owl | *Asio flammeus* |
| Scavenger / Bird of Prey | Long-eared Owl | *Asio otus* |
| Scavenger / Bird of Prey | Little Owl | *Athene noctua* |
| Scavenger / Bird of Prey | Bonaparte's Gull | *Larus philadelphia* |
| Scavenger / Bird of Prey | Spotted Eagle-Owl | *Bubo africanus* |
| Scavenger / Bird of Prey | Eurasian Eagle-Owl | *Bubo bubo* |
| Scavenger / Bird of Prey | Snowy Owl | *Bubo scandiacus* |
| Scavenger / Bird of Prey | Great Horned Owl | *Bubo virginianus* |
| Scavenger / Bird of Prey | Common Buzzard | *Buteo buteo* |
| Scavenger / Bird of Prey | Red-tailed Hawk | *Buteo jamaicensis* |
| Scavenger / Bird of Prey | Eastern Buzzard | *Buteo japonicus* |
| Scavenger / Bird of Prey | Rough-legged Hawk | *Buteo lagopus* |
| Scavenger / Bird of Prey | Red-shouldered Hawk | *Buteo lineatus* |
| Scavenger / Bird of Prey | Broad-winged Hawk | *Buteo platypterus* |
| Scavenger / Bird of Prey | Jackal Buzzard | *Buteo rufofuscus* |
| Scavenger / Bird of Prey | Swainson's Hawk | *Buteo swainsoni* |
| Scavenger / Bird of Prey | Northern Crested Caracara | *Caracara cheriway* |
| Scavenger / Bird of Prey | Turkey Vulture | *Cathartes aura* |
| Scavenger / Bird of Prey | Brown-headed Gull | *Chroicocephalus brunnicephalus* |
| Scavenger / Bird of Prey | Grey-headed Gull | *Chroicocephalus cirrocephalus* |
| Scavenger / Bird of Prey | Hartlaub's Gull | *Chroicocephalus hartlaubii* |
| Scavenger / Bird of Prey | Black-headed Gull | *Chroicocephalus ridibundus* |
| Scavenger / Bird of Prey | Western Marsh Harrier | *Circus aeruginosus* |
| Scavenger / Bird of Prey | Hen Harrier | *Circus cyaneus* |
| Scavenger / Bird of Prey | Northern Harrier | *Circus hudsonius* |
| Scavenger / Bird of Prey | Black Vulture | *Coragyps atratus* |
| Scavenger / Bird of Prey | Pied Crow | *Corvus albidae* |
| Scavenger / Bird of Prey | American Crow | *Corvus brachyrhynchos* |
| Scavenger / Bird of Prey | Common Raven | *Corvus corax* |
| Scavenger / Bird of Prey | Hooded Crow | *Corvus cornix* |
| Scavenger / Bird of Prey | Carrion Crow | *Corvus corone* |
| Scavenger / Bird of Prey | Rook | *Corvus frugilegus* |
| Scavenger / Bird of Prey | Large-billed Crow | *Corvus macrorhynchos* |
| Scavenger / Bird of Prey | Fish Crow | *Corvus ossifragus* |
| Scavenger / Bird of Prey | House Crow | *Corvus splendens* |
| Scavenger / Bird of Prey | Saker Falcon | *Falco cherrug* |
| Scavenger / Bird of Prey | Merlin | *Falco columbarius* |
| Scavenger / Bird of Prey | Prairie Falcon | *Falco mexicanus* |
| Scavenger / Bird of Prey | Peregrine Falcon | *Falco peregrinus* |
| Scavenger / Bird of Prey | Common Kestrel | *Falco tinnunculus* |
| Scavenger / Bird of Prey | Gyrfalcon | *Falco rusticolus* |
| Scavenger / Bird of Prey | American Kestrel | *Falco sparverius* |
| Scavenger / Bird of Prey | Red-footed Falcon | *Falco vespertinus* |
| Scavenger / Bird of Prey | Variable Hawk | *Geranoaetus polyosoma* |
| Scavenger / Bird of Prey | Bearded Vulture | *Gypaetus barbatus* |
| Scavenger / Bird of Prey | White-backed Vulture | *Gyps africanus* |
| Scavenger / Bird of Prey | Griffon Vulture | *Gyps fulvus* |
| Scavenger / Bird of Prey | American Oystercatcher | *Haematopus palliatus* |
| Scavenger / Bird of Prey | White-tailed Eagle | *Haliaeetus albicilla* |
| Scavenger / Bird of Prey | Bald Eagle | *Haliaeetus leucocephalus* |
| Scavenger / Bird of Prey | Steller's Sea Eagle | *Haliaeetus pelagicus* |
| Scavenger / Bird of Prey | African Fish Eagle | *Haliaeetus vocifer* |
| Scavenger / Bird of Prey | Bonelli's Eagle | *Hieraaetus fasciatus* |
| Scavenger / Bird of Prey | Pallas's Gull | *Ichthyaetus ichthyaetus* |
| Scavenger / Bird of Prey | Mediterranean Gull | *Ichthyaetus melanocephalus* |
| Scavenger / Bird of Prey | Inca Tern | *Larosterna inca* |
| Scavenger / Bird of Prey | Herring Gull | *Larus argentatus* |
| Scavenger / Bird of Prey | Armenian Gull | *Larus armenicus* |
| Scavenger / Bird of Prey | Short-billed Gull | *Larus brachyrhynchus* |
| Scavenger / Bird of Prey | Caspian Gull | *Larus cachinnans* |
| Scavenger / Bird of Prey | Belcher's Gull | *Larus belcheri* |
| Scavenger / Bird of Prey | California Gull | *Larus californicus* |
| Scavenger / Bird of Prey | Mew Gull | *Larus canus* |
| Scavenger / Bird of Prey | Ring-billed Gull | *Larus delawarensis* |
| Scavenger / Bird of Prey | Kelp Gull | *Larus dominicanus* |
| Scavenger / Bird of Prey | Lesser Black-backed Gull | *Larus fuscus* |
| Scavenger / Bird of Prey | Iceland Gull | *Larus glaucoides* |
| Scavenger / Bird of Prey | Great Black-backed Gull | *Larus marinus* |
| Scavenger / Bird of Prey | Yellow-legged Gull | *Larus michahellis* |
| Scavenger / Bird of Prey | Silver Gull | *Larus novaehollandiae* |
| Scavenger / Bird of Prey | Slaty-backed Gull | *Larus schistisagus* |
| Scavenger / Bird of Prey | Thayer's Gull | *Larus thayeri* |
| Scavenger / Bird of Prey | Franklin's Gull | *Leucophaeus pipixcan* |
| Scavenger / Bird of Prey | Eastern Screech-Owl | *Megascops asio* |
| Scavenger / Bird of Prey | Western Screech-Owl | *Megascops kennicottii* |
| Scavenger / Bird of Prey | Red Kite | *Milvus milvus* |
| Scavenger / Bird of Prey | Black Kite | *Milvus migrans* |
| Scavenger / Bird of Prey | Northern Gannet | *Morus bassanus* |
| Scavenger / Bird of Prey | Mountain Hawk-Eagle | *Nisaetus nipalensis* |
| Scavenger / Bird of Prey | Scops Owl | *Otus scops* |
| Scavenger / Bird of Prey | Ivory Gull | *Pagophila eburnean* |
| Scavenger / Bird of Prey | Osprey | *Pandion haliaetus* |
| Scavenger / Bird of Prey | Harris's Hawk | *Parabuteo unicinctus* |
| Scavenger / Bird of Prey | Black-billed Magpie | *Pica hudsonia* |
| Scavenger / Bird of Prey | Common Magpie | *Pica pica* |
| Scavenger / Bird of Prey | Secretary Bird | *Sagittarius serpentarius* |
| Scavenger / Bird of Prey | Crested Serpent Eagle | *Spilornis cheela* |
| Scavenger / Bird of Prey | Owl | *Strigiformes* |
| Scavenger / Bird of Prey | Tawny Owl | *Strix aluco* |
| Scavenger / Bird of Prey | Ural Owl | *Strix uralensis* |
| Scavenger / Bird of Prey | Barred Owl | *Strix varia* |
| Scavenger / Bird of Prey | Cape Gannet | *Sula capensis* |
| Scavenger / Bird of Prey | Elegant Tern | *Thalasseus elegans* |
| Scavenger / Bird of Prey | Common Barn-Owl | *Tyto alba* |
| Scavenger / Bird of Prey | Sabine's Gull | *Xema sabini* |

**Supplementary Figure S1.** Pairwise correlation heatmap of all candidate predictors.

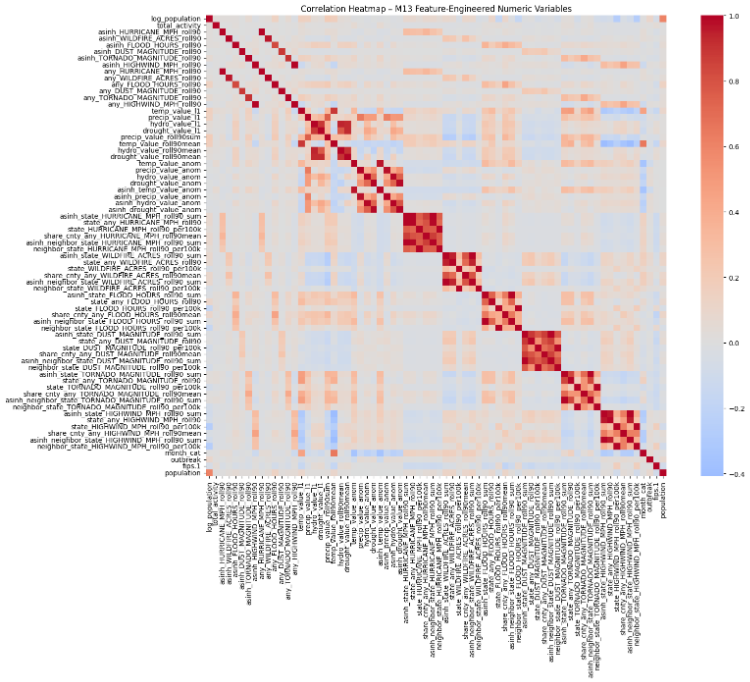
